# Contributions of rare and common variation to early-onset and atypical dementia risk

**DOI:** 10.1101/2023.02.06.23285383

**Authors:** Carter A. Wright, Jared W. Taylor, Meagan Cochran, James M.J. Lawlor, Belle A. Moyers, Michelle D. Amaral, Zachary T. Bonnstetter, Princess Carter, Veronika Solomon, Richard M. Myers, Marissa Natelson Love, David S. Geldmacher, Sara J. Cooper, Erik D. Roberson, J. Nicholas Cochran

## Abstract

We collected and analyzed genomic sequencing data from individuals with clinician-diagnosed early-onset or atypical dementia. Thirty-two patients were previously described, with sixty-eight newly described in this report. Of those sixty-eight, sixty-two patients reported Caucasian, non-Hispanic ethnicity and six reported as African American, non-Hispanic. Fifty-three percent of patients had a returnable variant. Five patients harbored a pathogenic variant as defined by the American College of Medical Genetics criteria for pathogenicity. A polygenic risk score was calculated for Alzheimer’s patients in the total cohort and compared to the scores of a late-onset Alzheimer’s cohort and a control set. Patients with early-onset Alzheimer’s had higher non-*APOE* polygenic risk scores than patients with late onset Alzheimer’s, supporting the conclusion that both rare and common genetic variation associate with early-onset neurodegenerative disease risk.

## Introduction

Dementia affects over 55 million people worldwide, nine percent of these patients are under age 65 (Dua et al. 2017). Rare variants in three genes, *PSEN1, PSEN2*, and *APP*, are associated with autosomal dominant early-onset Alzheimer’s disease (EOAD), however they only explain about 10% of genetic cases (Stoychev et al. 2019). The use of genome sequencing allows identification of rare variants not included in some targeted gene panel testing, as well as variation that is the purview of future research such as including non-coding variants and variants in novel risk loci. In our previous study we reported on the use of genome sequencing in 32 individuals with early-onset and/or atypical dementia and described several pathogenic variants associated with disease, including combinations of disease-associated risk variants. Our previous work confirmed the value of genetic assessment and identified contributing genetic variation for over half of the cohort (Cochran et al. 2019).

The American College of Medical Genetics (ACMG) criteria was used to assess pathogenicity and pathogenic/likely pathogenic variants were returned. Additional criteria to return variants included: 1) any variant with a disease-established odds ratio of >2 described in multiple reports, which we defined as an “established risk variant”, 2) presence of one or two *APOE ε4* alleles in a patient with EOAD or atypical dementia likely due to EOAD, or 3) one strong report with a disease-associated odds ratio >2 with replication included in the study design, which we defined as a “likely risk variant.”

In this report we also assessed the dementia risk related to common variation for the enrolled patients by calculating polygenic risk scores. Our results highlight the complex genetic etiology of early-onset and/or atypical dementia.

## Results

Here we report on 68 additional patients collected as a continuation of the previously published cohort. For each patient with clinician-diagnosed early-onset or atypical dementia, we collected and analyzed genomic sequencing data. We identified returnable, primary findings for 53% of patients **(Fig. 1A, Table 1)**. Including the initial 32 probands described by Cochran, et al. in our previous report, a total of 100 patients have been enrolled through the Brain Aging and Memory Clinic at the University of Alabama at Birmingham **(Fig. 1B)**.

**Figure 1.**
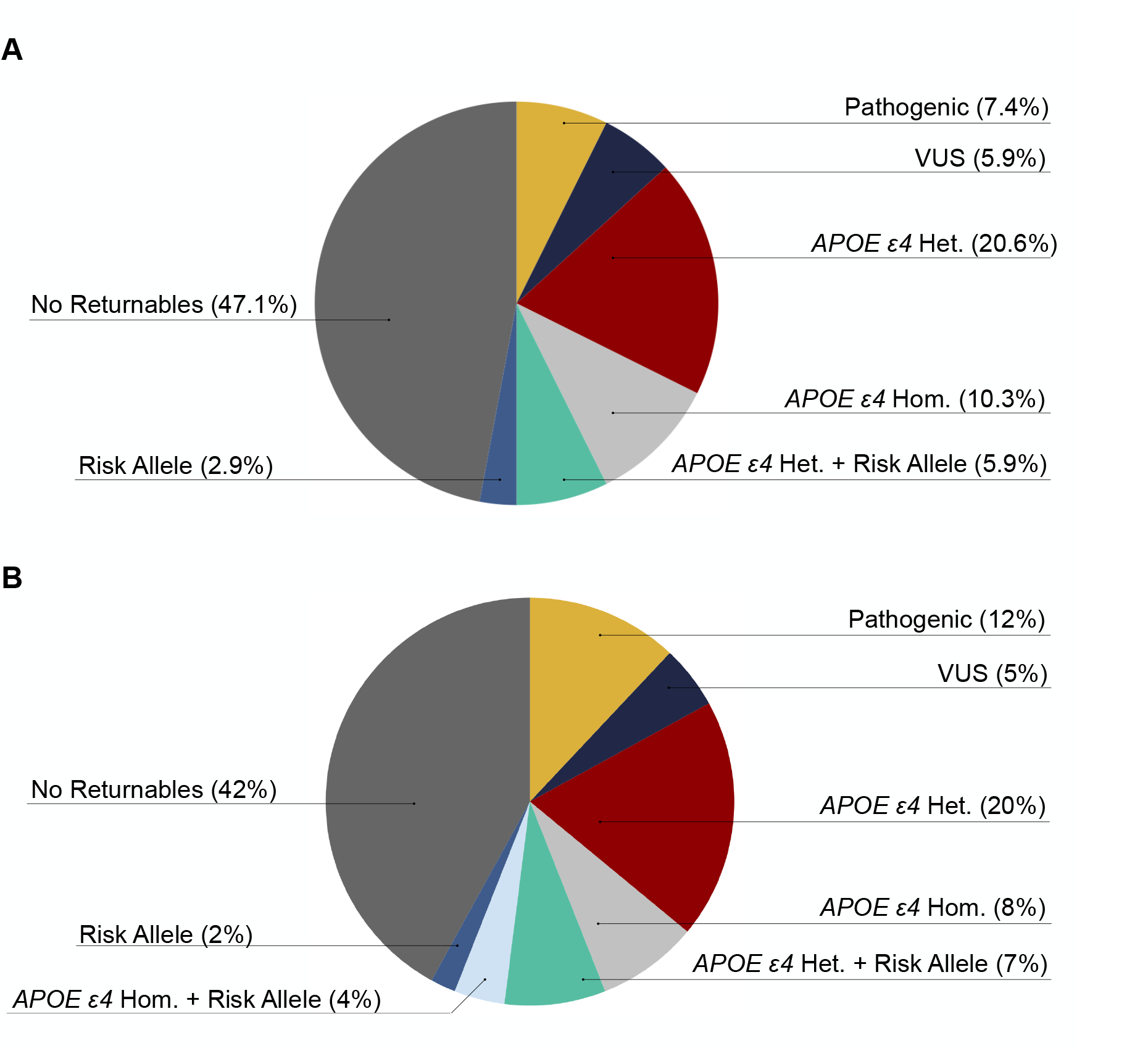
Summary of genomic sequencing findings for the (A) 68 proband cohort and (B) all 100 enrolled probands including the 32 described in Cochran, et al. Patients carrying one *APOE ɛ4* allele are noted as *APOE* ɛ4 Het. and those carrying two alleles are noted as *APOE ɛ4 H*om. Patients carrying a risk allele in addition to one or two *APOE ɛ4* allele are listed as *APOE ε4* Het. or Hom. + risk. VUS = Variant of Uncertain Significance.

**Table 1.** Variant Table.

| Gene | Chr. | HGVS DNA | HGVS Protein | Variant Type | Predicted Effect | dbSNP ID | CADD Score | gnomAD Allele Counts |
| --- | --- | --- | --- | --- | --- | --- | --- | --- |
| *APOE* | 19 | NM_000041.3:c.388T>C | p.(Cys130Arg) | SNV | Missense | rs429358 | 16.65 | 23,875 |
| *APOE* | 19 | NM_000041.3:c.526C>T | p.(Arg176Cys) | SNV | Missense | rs7412 | 26 | 31,094 |
| *SORL1* | 11 | NM_003105.6:c.3856C>T | p.(Arg1286Cys) | SNV | Missense | NA | 29.3 | 1 |
| *PRNP* | 20 | NM_183079.3:c.416T>C | p.(Ile139Thr) | SNV | Missense | NA | 18.98 | 0 |
| *SORL1* | 11 | NM_003105.6:c.6194A>T | p.(Asp2065Val) | SNV | Missense | NA | 28.8 | 347 |
| *ABCA7* | 19 | NM_019112.3:c.4416+1G>T | NA | SNV | Splice | NA | 26.3 | 6 |
| *PAH* | 12 | NM_000277.3:c.117C>G | p.(Phe39Leu) | SNV | Missense | rs62642926 | 25.6 | 13 |
| *PAH* | 12 | NM_000277.3:c.194T>C | p.(Ile65Thr) | SNV | Missense | rs75193786 | 27.2 | 45 |
| *GRN* | 17 | NM_002087.3:c.26C>A | p.(Ala9Asp) | SNV | Missense | rs63751243 | 23.6 | 0 |
| *MAPT* | 17 | NM_005910.5:c.1216C>T | p.(Arg406Trp) | SNV | Missense | rs63750424 | 23.5 | 4 |
| *PSEN1* | 14 | NM_000021.4:c.236C>T | p.(Ala79Val) | SNV | Missense | rs63749824 | 26.8 | 5 |
| *PSEN2* | 1 | NM_000447.3:c.712C>T | p.(Leu238Phe) | SNV | Missense | rs367855127 | 25.3 | 3 |
| *PSEN2* | 1 | NM_000447.3:c.211C>T | p.(Arg71Trp) | SNV | Missense | rs140501902 | 22.4 | 513 |
| *ABCA7* | 19 | NM_019112.3:c.4010C>T | p.(Thr1337Ile) | SNV | Missense | NA | 23.9 | 0 |
| *TSC2* | 16 | NM_000548.5:c.3846_3855del CTCCAAGGA | p.(Ser1282Arg_delC1283-G1285) | Deletion | INDEL | NA | NA | 0 |
| *SCN5A* | 3 | NM_198056.2:c.673C>T | p.(Arg225Trp) | SNV | Missense | rs199473072 | 23.4 | 12 |
| *KCTD17* | 22 | NM_001282684.2:c.299G>C | p.(Gly100Ala) | SNV | Missense | NA | 34 | 2 |
| *SORL1* | 11 | NM_003105.6:c.2711G>A | p.(Arg904Gln) | SNV | Missense | NA | 31 | 20 |

All variants were observed in the heterozygous state except for *APOE* (NM\_000041.3:c.388T>C, p.(Cys130Arg)), which was observed in both the heterozygous and homozygous state. *APOE* (NM\_000041.3:c.526C>T, p.(Arg176Cys)) is also noted here, because confirmation of its absence along with *APOE* (NM\_000041.3:c.388T>C, p.(Cys130Arg)) indicates the *APOE* ε4 allele. (HGVS) Human Genome Variation Society, (dbSNP) Single Nucleotide Polymorphism Database, (gnomAD) Genome Aggregation Database, (CADD) Combined Annotation Dependent Depletion, (SNV) single-nucleotide variant.

### Clinical Presentation and Family History

Of the 68 additional patients described here, 40 patients were male and 28 patients were female. Ethnicity was self-reported of which 62 patients reported Caucasian and 6 reported African American. All patients reported non-Hispanic ethnicity. One patient reported age of onset in the 80s, 4 in 70s, 23 in 60s, 29 in 50s, 10 in 40s, and one in 30s **(Supplemental Table 1)**. The nine patients with an age of onset older than 65 had moderate to strong family histories of dementia.

We generated a family history score for each patient that is a derivative of the scoring system first used by Goldman et al. (Goldman et al. 2005). The modified Goldman Score was generated as follows: (1) At least three people in two generations affected with EOAD, Frontotemporal dementia (FTD), ALS, corticobasal degeneration (CBD), Parkinson’s disease (PD), or progressive supranuclear palsy (PSP) with one person being a first-degree relative of the other two; (1.5) Criteria matching (1) but with late-onset Alzheimer disease (LOAD) instead of EOAD; (2) At least two relatives with dementia, FTD, ALS, CBD, PD, or PSP and criteria for autosomal dominant inheritance were not met; (3) A single affected first or second degree family member with early-onset dementia or at least two with FTD, ALS, CBD, PD, mild cognitive impairment (MCI), or PSP; (3.5) A single affected first or second degree family member with late-onset dementia, FTD, ALS, CBD, PD, MCI, or PSP; and (4) non contributory family history or unknown family history. We classified patients with a family history score of 1 or 1.5 as strong family history, 2, 3, or 3.5 as moderate family history, and those with a score of 4 had no contributory or known family history. All family history scores are included in **Supplemental Table 1**. We note that the researchers do not have the ability to identify patients based on IDs, as the key is kept at the clinical enrollment site.

#### Genomic Analyses

Variants were evaluated using ACMG criteria (Richards et al. 2015) for pathogenicity and the ACMG evidence codes for all pathogenic variants are detailed in the **Supplemental ACMG Pathogenicity Evidence Details**.

### C9orf72 Expansion Testing

We tested for pathogenic G_4_C_2_ hexanucleotide expansion at the *C9orf72* locus, which is associated with FTD and ALS, either as described in Cochran, et al. (Cochran et al. 2019) (using a separately obtained test from GeneDx) or, for samples that were sequenced with a PCR-free genome, using ExpansionHunter (Dolzhenko et al. 2017). All patients in this cohort expansion of 68 were negative for *C9orf72* repeat expansion.

#### Pathogenic Diagnoses

### Two Pathogenic Variants in PAH in a Patient with Phenylketonuria

A patient with phenylketonuria (PKU) with onset in their early 40s had two pathogenic variants in *PAH* (NM_000277.3:c.117C>G, p.(Phe39Leu)) and (NM_000277.3:c.194T>C, p.(Ile65Thr)). Segregation analysis of the family showed the *PAH* variants were in *trans*. Both variants have been submitted to ClinVar as pathogenic for PKU, by multiple submitters (VCV000000605.15 and VCV000000636.18). PKU is an autosomal recessive disease that can cause brain dysfunction due to increased phenylalanine concentrations (van Spronsen et al. 2021). Adult-onset PKU is rare and can present with symptoms of dementia and parkinsonism making it difficult to quickly diagnose. If diagnosed correctly PKU can be treated with a phenylalanine restricted diet (Tufekcioglu et al. 2016; Rosini et al. 2014).

### A Pathogenic GRN Variant in a Family with FTD

A patient with an onset of MCI in their early 50s had a pathogenic variant in *GRN* (NM_002087.3:c.26C>A, p.(Ala9Asp)). This variant is absent from gnomAD and has been reported by several laboratories as pathogenic in ClinVar (VCV000016013.12). This variant segregates with two affected family members. We classified this variant as pathogenic as additional studies have shown this variant to segregate with FTD (Mukherjee et al. 2008; Shankaran et al. 2008).

### A Rare MAPT Variant in Five Early-onset Alzheimer’s Disease Patients

Two patients with EOAD (one patient also exhibited signs of FTD) with age of onset in their 50s have a pathogenic variant in MAPT (NM_005910.5:c.1216C > T, p.(Arg406Trp)). This rare variant has a minor allele frequency of 0.000016% and was reported by several laboratories as pathogenic in ClinVar (VCV000014247.18). In our previous study (Cochran et al. 2019), we identified three patients with this variant in *MAPT*. The presence of the same rare variant in a total of five patients enrolled at the same clinic suggests a common ancestry is likely **(Fig. 2A)**. We inferred the relatedness of patients using KING (Manichaikul et al. 2010) and determined a kinship coefficient for each pair of relationships and determined that three probands have a kinship coefficient of 0.06 or greater, reaching the criteria for third degree relation **(Fig. 2B)**. The other values did not reach the kinship coefficient of 0.06 criteria but were near that threshold and might reflect a more distant relationship.

**Figure 2.**
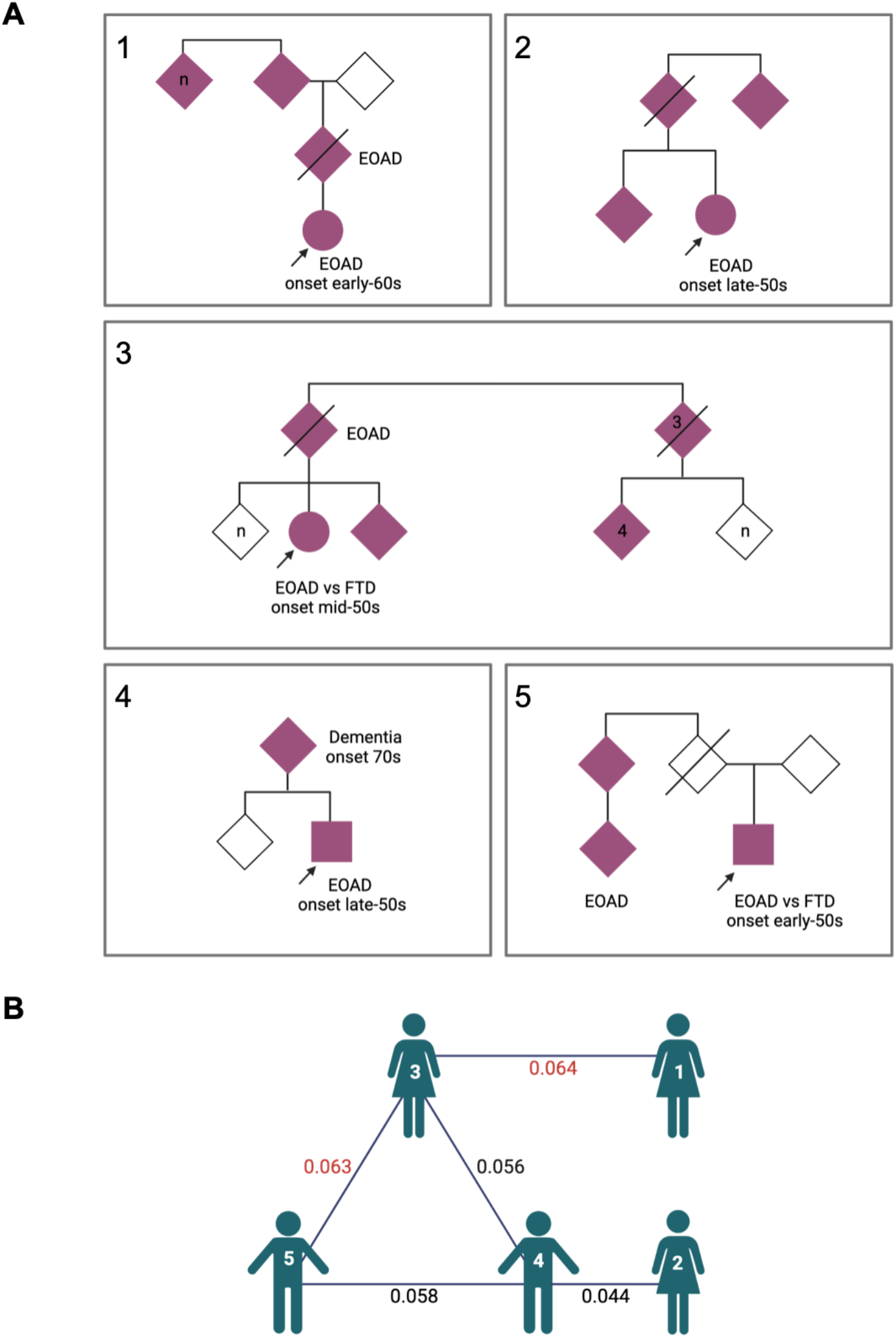
Evaluation of five probands with a *MAPT* R406W variant. (A) Pedigrees for the five probands with the *MAPT* variant. Probands are marked with arrow. Affected family members are solid pink. Patients 1-3 were identified in our previous study (Cochran et al., 2019). (B) Diagram showing the KING kinship coefficients between probands. Values in red denote third degree relatives. Figure was generated at biorender.com.

### A PSEN1 Variant in a Patient with Mild Cognitive Impairment

An individual with symptoms of MCI and a strong family history of AD had a pathogenic variant in *PSEN1* (NM_000021.4:c.236C>T, p.(Ala79Val)). Onset of symptoms began in their 60s and multiple affected family members had onset in 60s. This pathogenic variant has been previously reported in ClinVar (VCV000018157.5) in patients with AD. Variants in *PSEN1* are the most common cause of autosomal dominant familial Alzheimer’s disease (Kelleher and Shen 2017).

#### Compound Heterozygous Risk Factors

### APOE ε4 Risk Factors

The *APOE ɛ4* allele is the most common strong risk factor for Alzheimer’s disease (AD), the most common form of dementia. Patients carrying one *APOE ɛ4* allele are noted as *APOE* ɛ4 heterozygous and those carrying two alleles are noted as *APOE ɛ4* homozygous. We identified 25 (36%) patients with at least one *APOE ɛ4* allele. Several of them, described here, also carried other risk variants.

### APOE ε4 with SORL1 Risk Variants

A patient presenting with symptoms of EOAD with onset in their early 50s and a strong family history of AD had one *APOE* ε4 allele, a “likely risk variant” in *SORL1* (NM_003105.6:c.3856C>T, p.(Arg1286Cys)), and a variant in *PRNP* (NM_183079.3:c.416T>C, p.(lle139Thr)) (returned as a VUS). Pathogenic *PRNP* variants are typically missense variants and are associated with prion diseases (Piazza et al. 2020). To our knowledge the c.416T>C variant in *PRNP* has not been previously published or reported in gnomAD (Karczewski et al. 2020). This variant was returned as a VUS because the family history of a prion disease could not be determined. The c.3856C>T *SORL1* variant has a minor allele frequency of 0.0007% and is predicted damaging by PolyPhen-2 (Adzhubei et al. 2010) and SIFT (Ng and Henikoff 2003), with a Combined Annotation Dependent Depletion (CADD) (Kircher et al. 2014) score of 29. Loss-of-function *SORL1* variant carriers are present at an odds ratio of ∼4 compared to population databases (Raghavan et al. 2018). The odds ratio for LOF variants in this gene could be as high as 12.3 for AD and 27.5 for EOAD. For rare missense variants, the odds ratio could be as high as 3.14 for EOAD (Campion et al. 2019). Other similar variants in *SORL1*, in aggregate, are enriched in patients with early-onset dementia by at least 2 fold in comparison to individuals without dementia (Holstege et al. 2017).

Two patients in our study harbored one *APOE* ε4 allele and the same “likely risk variant” in *SORL1* (NM_003105.6:c.6194A>T, p.Asp2065Val). This variant in the *SORL1* gene has not been extensively described, to our knowledge. One of these two patients was diagnosed with EOAD, with an onset in their late 40s. The other enrolled patient harboring this variant presented with mild cognitive impairment that was amnestic in character with an age of onset in their 70s and a striking family history of dementia. This patient also had a rare variant in *ECE2* (NM_001037324.2:c.1120C>T, p.(Arg374*)) that was not returned but had a CADD score of 39 and had a predicted effect of a stop gained. Variants in *ECE2* have been implicated in Alzheimer’s disease risk, and variants in the peptidase domain were shown to impair enzymatic activity of ECE2 in Aβ degradation (Liao et al. 2020).

### APOE ε4 Heterozygote with ABCA7 Risk Allele

An African American patient diagnosed with mild cognitive impairment with onset in their late 50s harbored an “established risk” loss-of-function splice variant in *ABCA7* (NM_019112.3:c.4416+1G>T) and one *APOE* ε4 risk allele. The *ABCA7* splice variant had a CADD score of 26 and is predicted to affect splicing by multiple splicing prediction tools (Pashaei et al. 2016; Jian et al. 2014). The c.4416+1G>T variant has been identified in gnomAD 6 times, however it has only been identified in African Americans. Variants in *ABCA7* have been implicated as strong genetic predictors of AD in African Americans (Reitz et al. 2013) and have been associated with the progression and development of AD (Sinha et al. 2019). These variants likely contribute to the individual’s strong family history of AD.

#### Rare Variants of Uncertain Significance

In a patient with onset in their 50s and a EOAD diagnosis, we observed a VUS in *PSEN2* (NM_000447.3:c.712C>T, p.(Leu238Phe)). This variant has a CADD of 25, is predicted damaging by PolyPhen-2 and MetaSVM (Kim et al. 2017), and has been previously classified as a VUS and once as likely pathogenic in Clinvar (VCV000448151.4). Hsu, et al. described this *PSEN2* L238F variant as a potential AD risk factor after identifying it in a patient diagnosed with EOAD that had several affected family members (Hsu et al. 2018). In another study, mouse neuroblastoma cells with this L238F variant had an increased Aβ42/40 ratio compared to wildtype cells which led to the classification for this variant as “probable pathogenic” (Hsu et al. 2020). An increased Aβ42/40 ratio is a predictor of amyloid-PET positivity in patients with Alzheimer’s disease (Amft et al. 2022).

A patient with EOAD had a variant in *PSEN2* (NM_000447.3:c.211C>T, p.(Arg71Trp)) (returned as “likely risk”). Previous studies reported the odds ratio for this variant as 6.45 and 10.3 (Benitez et al. 2013; Cruchaga et al. 2012), suggesting that someone who has the variant is at increased risk to develop AD, but these reports suggest this variant alone is not enough to cause disease. Additionally, individuals who have AD and carry this variant have a significantly earlier age of onset than those who have AD and do not carry the variant (Cruchaga et al. 2012). This is consistent with our finding of EOAD in this patient.

One individual with AD with age of onset in their 50s had a rare variant in *ABCA7* (NM_019112.3:c.4010C>T, p.(Thr117Ile)) (returned as a VUS) that has not been identified in gnomAD or published to our knowledge. Rare missense variants in *ABCA7* have been previously reported to have an odds ratio of 1.6 and loss-of-function variants have an odds ratio of 2.2 for individuals with EOAD (Holstege et al. 2021). This individual met the missense criteria (allele frequency <1% and ‘moderate’ or ‘high’ variant effect predictor impact classification) described by Holstege, et al. We predict this rare variant to be damaging due to its CADD score (23.9) and computational evidence supporting its deleterious effect on the gene or gene product.

An individual diagnosed with moderate dementia with age of onset in their 60s had a rare deletion in *TSC2* (NM_000548.5:c.3846_3855delCTCCAAGGA, p.(Ser1282Arg_delC1283–G1285)) (returned as a VUS). This individual also had a secondary finding: a pathogenic variant in *SCN5A*(NM_000335.4:c.673C>T, p.(Arg225XTrp)). This specific *SCN5A* variant has been reported to cause Brugada syndrome (Beckermann et al. 2014; Kapplinger et al. 2010). Brugada syndrome is an inherited cardiac arrhythmia condition and management includes prevention of cardiac arrest. The individual’s sibling was reported to have died from cardiac arrest in their 60s.

A patient diagnosed with EOAD with age of onset in their 40s had a rare variant in *KCTD17* (NM_001282684.2:c.299G>C, p.(Gly100Ala)) (returned as a VUS). This variant had a CADD of 34 and was predicted damaging due to the computational evidence supporting its deleterious effect on the gene or gene product. The c.299G>C variant in the *KCTD17* gene was previously classified as a VUS for Myoclonus-dystonia by Invitae (Accession number SCV000932534.2 in ClinVar).

A patient diagnosed with moderate dementia with age of onset in the 50s had a rare variant in *SORL1* (NM_003105.6:c.2711G>A, p.(Arg904Gln)) (returned as “likely risk”). As previously described above, *SORL1* missense variants with a minor allele frequency < 1×10^−4^ and multiple bioinformatic damaging predictions have a corresponding odds ratio of ∼3 (Campion et al. 2019). This variant had a CADD of 31 and was predicted damaging by PolyPhen-2, MetaSVM, and PROVEAN (Choi and Chan 2015).

#### Polygenic Risk Score

A polygenic risk score (PRS) represents a statistical estimate of disease risk calculated by combining information about multiple risk variants throughout the genome. We sought to understand the contribution of common variation to AD risk in this cohort.

Using the PRS generated by Cruchaga, et al. (Cruchaga et al. 2018), we computed a PRS for each proband in our study **(Supplemental Table 2)**. We also calculated scores for 880 LOAD cases (818 ADSP, 62 UAB), 77 EOAD (UAB early-onset or atypical dementia) cases that are the focus of this study, and 5,179 healthy controls (5,043 ADSP, 136 other Alabama-based study). We calculated PRS for all individuals using two methods, one that incorporates *APOE* status and one that omits it. *APOE* is a strong known risk factor for AD, however when including it in the PRS there was not a significant different between dementia cohorts **(Supplemental Fig. 1)**. We did observe differences in the PRS of these cohorts when *APOE* was excluded (**Fig. 3**). We observed that the median PRS of the UAB EOAD cohort was higher than the LOAD cohort upon *APOE* exclusion. These results are consistent with the possibility that *APOE* status alone could be a stronger association for LOAD; in contrast, though *APOE* remains a critical contributor to EOAD, additional deleterious genetic variation may be more likely to contribute to the emergence of an EOAD presentation. We also showed that the PRS in all AD cohorts was higher than our control cohort of unaffected individuals **(Fig. 3)**. Since we combined two control cohorts and two LOAD cohorts we also compared the PRS of each cohort separately and did not observe any significant differences between control or LOAD groups **(Supplemental Fig. 2**). In the 42% of UAB EOAD patients with no returnable findings, we hypothesized that PRS might be higher than those patients with a known pathogenic variant due to the combination of multiple genetic factors contributing to dementia risk. That would indicate that perhaps common variation rather than rare variation were contributing to risk in these cases. However, in the UAB EOAD cohort we did not see a significant difference in PRS between patients with a returnable pathogenic variant based on genomic sequencing compared to those with no returnable variants **(Supplemental Fig. 3)**.

**Figure 3.**
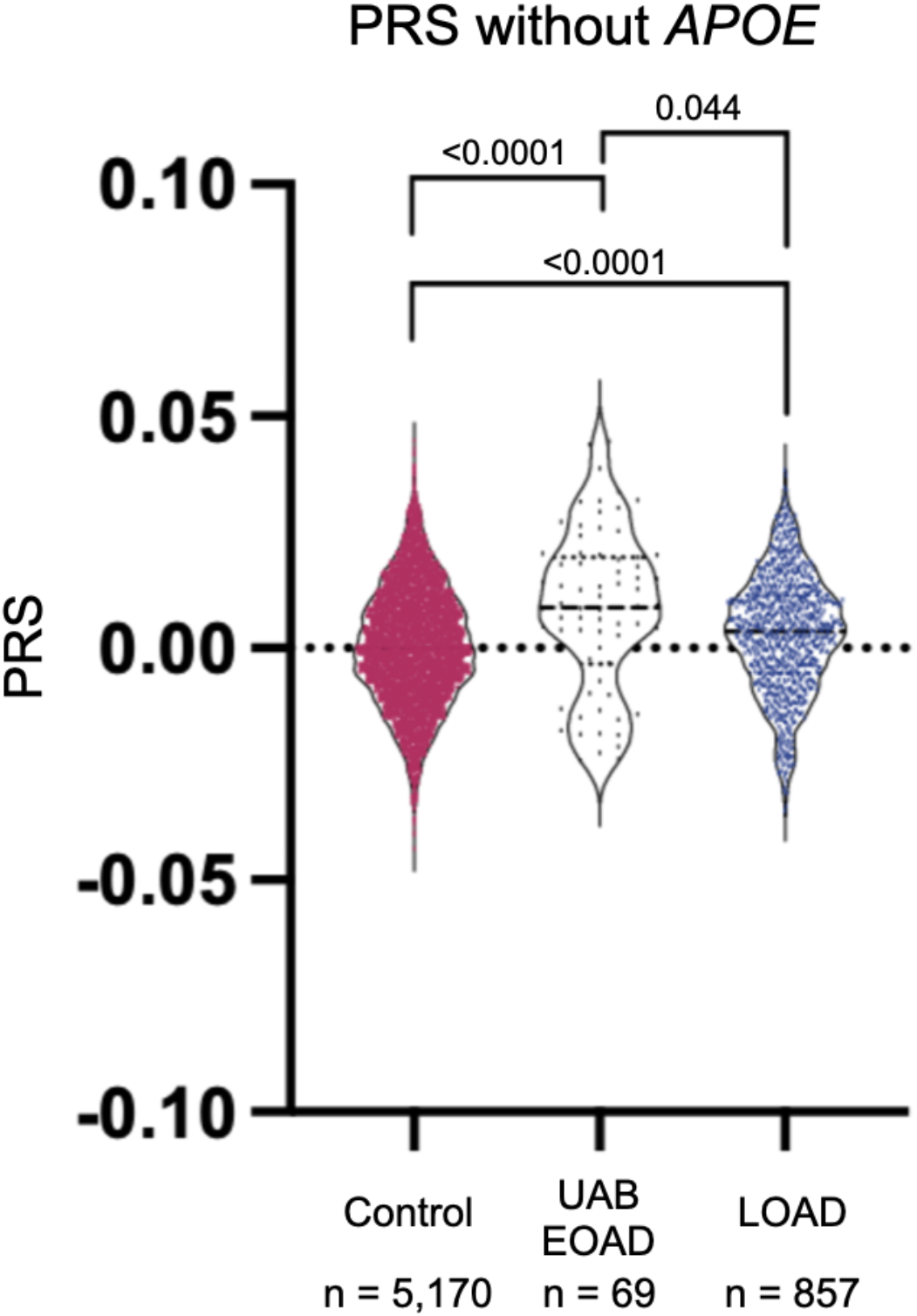
Violin plot comparing polygenic risk scores (PRS) of the UAB early-onset Alzheimer’s (UAB EOAD) cohort to patients with a late-onset Alzheimer’s (LOAD) cohort, and the control (Control) cohort. This PRS excludes *APOE* status. Only individuals with a self-reported ancestry of Caucasian, Non-Hispanic were included in this analysis. ANOVA with Kruskal-Wallis test was used to calculate adjusted p-values.

Individuals with the strongest family history are more likely to have a returnable result (regardless of PRS), but 71% of non-Hispanic Caucasian patients with the strongest family history of early-onset or atypical dementia still do not have a known pathogenic mutation. This led us to consider if combinations of common variants can explain risk in these individuals. Indeed, there is a correlation where a more extensive family history of dementia is associated with significantly higher *APOE*– PRS, but there is no effect on *APOE*+ PRS **(Supplemental Fig. 4)**.

African Americans have a higher prevalence of AD than Caucasians, however there are disparities in the amount of genetic data available from this population (Clark et al. 2022). The generation of polygenic risk scores are typically performed using Caucasian, non-Hispanic populations. As others have observed (Clark et al. 2022), we saw that existing PRS do not capture variation contributing to AD risk in African Americans **(Supplemental Fig. 5)**, pointing to the importance of developing ancestry-specific and/or ancestry adjusted PRS.

## Discussion

Accumulation of amyloid-β protein in the brain is characteristic of Alzheimer’s disease and can occur many years before the onset of symptoms (Luckett et al. 2022). As a prominent example, *APOE*, the largest genetic risk factor for AD, has been associated with amyloid-β accumulation in the brain, therefore identifying genetic risk variants prior to symptom onset is critical for treatment decisions and potential enrollment in prevention trials. In this study, genome sequencing revealed variants contributing to dementia risk that would be missed by targeted panel testing, affirming the need for comprehensive genetic approaches. For example, *MAPT* variants are typically associated with FTD, however there have been multiple reports of *MAPT* variants in patients with AD (Cruts et al. 2012; Reed et al. 1997; Rademakers et al. 2003). Panels that are specific to AD do not include *MAPT* variants, thus this variant would not be detected. Whole genome sequencing provides a solution to this challenge. Some commercially available panels address this issue by assessing genes associated with multiple neurodegenerative diseases on the same panel. Six percent of our 100 patient cohort had variation in *MAPT* that contributes to AD risk, however in this cohort, we hypothesized that the five patients with the *MAPT* R406W variant likely shared common ancestry limiting the interpretation of the frequency of mutations in this gene leading to AD risk.

To better assess the risk of dementia patients with multiple contributory factors we calculated a PRS for individuals with early-onset or atypical dementia that was either definitively or most likely due to AD and late-onset AD, as well as a control cohort. The PRS we used was employed in a previous study (Cruchaga et al. 2018). Similar to Cruchaga, et al. we showed an increased PRS in patients with an earlier onset of disease when considering *APOE*– PRS, however we observed the opposite for *APOE*+ PRS. This points to the importance of further studies to clarify the applicability of LOAD PRS to EOAD, and suggests that EOAD-specific PRS could be informative as EOAD cohort sizes grow to a size that permits generation of a reliable EOAD-specific score.

This trend held true when separating cohorts by ethnicity, however when applied to African American samples the PRS were lower compared to Caucasian samples with similar phenotypes and in fact African American patients with LOAD did not have a significantly different PRS than the controls. African Americans have twice the risk of developing AD as Caucasian individuals (Alzheimer’s disease facts and figures, 2020). Future studies generating PRS using genetic data from African Americans and other non-Europeans is necessary to more accurately determine the contribution of common variation in African American individuals, but the latest GWAS for African Americans with AD is still underpowered for this analysis, pointing to the critical nature of further study in this population (Kunkle et al. 2021).

In this study, we showed that multiple genetic factors, both rare and common, contribute to an individual’s dementia risk. Variants were not returned to many patients and this could be because our cohort is limited by patient number, thus it is possible that a larger cohort would reveal small effects of PRS in patients with no returnable findings. Despite the limitation of patient number, we identified and returned variants that contribute to the genetic explanation of patients’ symptoms for more than half of the cohort. This study contributes to the body of work showing the value of comprehensive genetic testing in identifying variants contributing to early-onset dementia risk (Cochran et al. 2019; Huq et al. 2022) and highlights that both rare and common genetic variation can associate with early-onset and/or atypical dementia.

## Methods

### *C9orf72* Expansion Testing

ExpansionHunter (Dolzhenko et al. 2019) was used to detect repeat expansions of *C9orf72* for samples with PCR-free genomes (21 individuals). Prior to implementation of PCR-free genomes, *C9orf72* expansions were assessed using a separate clinical test (GeneDx).

### Polygenic Risk Score

PRS was calculated with and without *APOE* using PLINK 1.9 --score with the no-mean-imputation option. Odds ratios for the SNPs were collected from the IGAP study (Lambert et al. 2013), and a log base 2 transformation was used on the odds ratios. *APOE* risk was calculated through the use of artificial SNPs such that each SNP represented the combination of rs429358 and rs7412 alleles. The odds ratios for the PRS scores with *APOE* used odds ratios from Farrer et al.(Farrer et al. 1997) for *APOE* status (ε3/ε4, ε4/ε4, ε2/ε3, ε2/ε4, ε2/ε2) instead of each SNP independently.

The HudsonAlpha CSER study enrolled and sequenced children with an early-onset neurodevelopmental disorder (NDD) with symptoms such as intellectual disability, seizures, developmental delays, etc. Many of these children’s parents were also sequenced, and we used these parents as controls. While these individuals were ascertained as a result of having a child with an NDD, the parents themselves were generally healthy. In that context, it is important to note that the vast majority of the disease-associated variation found in the CSER study was either de novo (i.e., the disease-associated allele in the proband is absent from his/her parents), recessive (i.e., each parent was a heterozygous, unaffected carrier of a disease-associated allele), or X-linked (i.e., the proband is hemizygous for a disease-associated allele inherited from an unaffected heterozygous mother). To the extent that some of these parental individuals harbor symptoms and/or dominant or incompletely penetrant risk factors of NDDs, such conditions are phenotypically and genetically distinct from the neurodegenerative diseases we are studying here. Finally, we note that a small amount of phenotypic data was collected for each of these parents, and none of them were known to have a neurodegenerative disease at enrollment.

Patients with a clinical diagnosis of FTD, CAA, MSA, PCA, CBS, white matter disease, progressive spastic dysarthria, or MS were excluded for polygenic risk score calculation.

### Genome Sequencing

Genome sequencing was performed at the HudsonAlpha Institute for Biotechnology on the NovaSeq platform using paired end 150-base pair reads. Sequencing libraries were prepared by Covaris shearing, end repair, adapter ligation, and PCR using standard protocols. Library concentrations were normalized using KAPA qPCR prior to sequencing. All sequencing variants returned to patients were validated by Sanger in a CAP/CLIA laboratory.

### Data Processing and Quality Control

Quality control included confirmation of each sample’s expected biological sex based on counts of chrX heterozygous variants. All samples were processed through a unified sequence alignment and variant calling pipeline. Variants were called with Strelka version 2.9.10. Sequence reads were aligned to GRCh38.p12 (with HudsonAlpha Clinical Sequencing Lab customized ALT mappings) using the Sentieon v201808.07 (Kendig et al. 2019) implementation of BWA-MEM (Li 2013) and command line option - M -K 10000000. BWAKit was used for post-alt processing of the alternate contig alignments. Duplicate reads were marked and base quality scores were recalibrated with Sentieon v201808.07 using dbSNP v.146, Mills, and 1000G gold standard indels as training data. Variants were called on the hg38 primary contigs (chr1-chr22, chrX, chrY, chrM) using Strelka v2.9.10 (Kim et al. 2018) in germline single-sample analysis mode.

### Data Analysis

Statistical analyses were conducted in R (version 3.6.1) using ggplot2 (version 3.3.6) (Wickham 2016).

Vcftools (version 0.1.16) (Danecek et al. 2011) was used for relationship inference analyses using the --relatedness2 command line option.

GraphPad Prism 9 (version 9.3.1) was used for plotting violin plots and ANOVA with Kruskal-Wallis tests performed in GraphPad were unpaired, non-parametric, two-tailed with 95% confidence interval.

### Genomic Analysis

The HudsonAlpha-developed Codicem application was used to analyze and support the interpretation of the variant data (described elsewhere (Holt et al. 2019)). Simple filtering for population allele frequencies (i.e., gnomAD and TOPMed Bravo (NHLBI 2018)), *in silico* deleteriousness scores (i.e., CADD, PolyPhen-2, and SIFT), and gene lists relevant to the phenotype of interest would recapitulate our findings using any suitable software package or even by a command line interface. In addition to searching for single-nucleotide variants and small indels, we also searched for large copy-number variations using four callers (DELLY (Rausch et al. 2012), ERDS (Zhu et al. 2012), CNVnator (Abyzov et al. 2011), and BIC-seq2 (Xi et al. 2016)), but did not identify any relevant to patient phenotypes (including absence of APP duplications).

### Return of Results

Results meeting criteria for return were delivered to patients by clinicians in the UAB Brain Aging and Memory Clinic through letters written by a genetic counselor. Letters included information on the variant, associated disease, recurrence risk, and management recommendations. Patients were given the option to have a genetic counselor present for return of results via phone or videoconference or to follow up with a genetic counselor after delivery of results. Primary results were provided only to probands. Although a secondary result was identified in only one participant who was a patient, we did also offer nonpatient participants (family members) receipt of actionable secondary findings (ACMG SF v3.0) if such a result had been identified. Family members of patients that received diagnostic results were provided with information to seek out clinical genetic counseling and targeted testing for familial variants if they desired.

Patients were able to opt in to receiving secondary findings. To return secondary findings we followed the ACMG classification criteria. Secondary variants had to reach a classification of likely pathogenic or pathogenic and must be in a gene on the ACMG SF v3.0 list.

## Additional Information

### Data Access

Data from the first 32 participants enrolled in this study are deposited at NIAGADS under project NG00082 - UAB/HudsonAlpha Families with Neurodegenerative Diseases. **Note to reviewers:** data for all other participants as well as for participants in the UAB ADRC study (a subset of late-onset cases) will be deposited to NIAGADS and made publicly available as a prerequisite of acceptance of the manuscript. We are working on this data deposition currently.

We combined two cohorts of LOAD patients. One LOAD cohort was collected at UAB and a Global Diversity (Illumina product #20031816) plus NeuroBooster microarray was run on the samples. The other LOAD samples were from the Alzheimer’s Disease Sequencing Project (ADSP) (NIAGIDS accession number: NG00067.v9). Control samples were from the HudsonAlpha CSER study (Bowling et al.), dbGap study accession number: phs001089.v3.p1) and from ADSP (NIAGIDS accession number: NG00067.v9).

## Supporting information

Supplemental ACMG Pathogenicity Evidence Details and Extended Acknowledgements

Publication License

## Data Availability

Data from the first 32 participants enrolled in this study are deposited at NIAGADS under project NG00082 - UAB/HudsonAlpha Families with Neurodegenerative Diseases. Note to reviewers: data for all other participants as well as for participants in the UAB ADRC study (a subset of late-onset cases) will be deposited to NIAGADS and made publicly available.
Control sample data is available from HudsonAlpha CSER study(dbGap study accession number: phs001089.v3.p1) and from ADSP (NIAGIDS accession number: NG00067.v9).

## Ethics Statement

This study was approved by UAB IRB protocol X161202004, “Evaluation of Genomic Variants in Patients with Neurologic Diseases.” All participants described provided explicit written consent for publication. Participants in the UAB LOAD set were approved via UAB IRB protocol #300000169, “UAB Alzheimer’s Disease Research Center/ Brain Aging and Memory in the South (BAMS) Study.”

## Acknowledgements

We thank the Clinical Services Laboratory at HudsonAlpha for DNA isolations, library generation, quality control, and sequencing and the Codicem software development team at HudsonAlpha for genome analysis software. We thank Dr. Emily Gordon for her assistance with pedigree visualization. We also thank the Greg Cooper Lab, including Drs. Michelle Thompson, Susan Hiatt, and Don Latner, for helpful discussions about variant interpretation.

## Authors’ Contributions

J.N.C. and E.D.R. designed the study. J.N.C., E.D.R., and R.M.M. secured funding. J.N.C. and E.D.R. wrote the IRB protocol. V.S. and P.C. coordinated all aspects of patient interaction. J.M.J.L., Z.T.B., and J.W.T. processed sequencing data. C.A.W., J.N.C., M.A., and B.A.M. analyzed genomes. C.A.W. coordinated genomic sequencing and led variant review committee meetings. C.A.W. and J.W.T. conducted other analyses. M.C. wrote clinical letters and provided genetic counseling. E.D.R., D.S.G. and M.N.L. recruited participants and returned results. C.A.W. wrote the manuscript, with edits by S.J.C. and J.N.C. All authors approved the final manuscript.

### Funding

Funding for genomes sequenced at HudsonAlpha was generously provided by the Daniel Foundation of Alabama and donors to the HudsonAlpha Foundation Memory and Mobility Program. Additional funding was provided by NIH grant R00AG068271 (J.N.C.), and NIH grant 5P20AG068024 (E.D.R., D.S.G., M.N.L, J.N.C., and J.W.T.).

## Notes

### Competing Interest Statement

The authors have declared no competing interest.

### Author Declarations

IRB of University of Alabama at Birmingham gave ethical approval for tis work.

## References

Abyzov A, Urban AE, Snyder M, Gerstein M. 2011. CNVnator: an approach to discover, genotype, and characterize typical and atypical CNVs from family and population genome sequencing. Genome Res 21: 974–984.

Adzhubei IA, Schmidt S, Peshkin L, Ramensky VE, Gerasimova A, Bork P, Kondrashov AS, Sunyaev SR. 2010. A method and server for predicting damaging missense mutations. Nat Methods 7: 248–249.

Amft M, Ortner M, Eichenlaub U, Goldhardt O, Diehl-Schmid J, Hedderich DM, Yakushev I, Grimmer T. 2022. The cerebrospinal fluid biomarker ratio Aβ42/40 identifies amyloid positron emission tomography positivity better than Aβ42 alone in a heterogeneous memory clinic cohort. Alzheimers Res Ther 14: 60.

Beckermann TM, McLeod K, Murday V, Potet F, George AL Jr. 2014. Novel SCN5A mutation in amiodarone-responsive multifocal ventricular ectopy-associated cardiomyopathy. Heart Rhythm 11: 1446–1453.

Benitez BA, Karch CM, Cai Y, Jin SC, Cooper B, Carrell D, Bertelsen S, Chibnik L, Schneider JA, Bennett DA, et al. 2013. The PSEN1, p.E318G variant increases the risk of Alzheimer’s disease in APOE-ε4 carriers. PLoS Genet 9: e1003685.

Bowling KM, Thompson ML, Amaral MD, Finnila CR, Hiatt SM, Engel KL, Nicholas Cochran J, Brothers KB, East KM, Gray DE, et al. 2017. Genomic diagnosis for children with intellectual disability and/or developmental delay. Genome Med. 9: 43

Campion D, Charbonnier C, Nicolas G. 2019. SORL1 genetic variants and Alzheimer disease risk: a literature review and meta-analysis of sequencing data. Acta Neuropathol 138: 173–186.

Choi Y, Chan AP. 2015. PROVEAN web server: a tool to predict the functional effect of amino acid substitutions and indels. Bioinformatics 31: 2745–2747.

Clark K, Leung YY, Lee W-P, Voight B, Wang L-S. 2022. Polygenic risk scores in Alzheimer’s disease genetics: Methodology, applications, inclusion, and diversity. J Alzheimers Dis 89: 1–12.

Cochran JN, McKinley EC, Cochran M, Amaral MD, Moyers BA, Lasseigne BN, Gray DE, Lawlor JMJ, Prokop JW, Geier EG, et al. 2019. Genome sequencing for early-onset or atypical dementia: high diagnostic yield and frequent observation of multiple contributory alleles. Cold Spring Harb Mol Case Stud 5. http://dx.doi.org/10.1101/mcs.a003491.

Cruchaga C, Del-Aguila JL, Saef B, Black K, Fernandez MV, Budde J, Ibanez L, Deming Y, Kapoor M, Tosto G, et al. 2018. Polygenic risk score of sporadic late-onset Alzheimer’s disease reveals a shared architecture with the familial and early-onset forms. Alzheimers Dement 14: 205–214.

Cruchaga C, Haller G, Chakraverty S, Mayo K, Vallania FLM, Mitra RD, Faber K, Williamson J, Bird T, Diaz-Arrastia R, et al. 2012. Rare variants in APP, PSEN1 and PSEN2 increase risk for AD in late-onset Alzheimer’s disease families. PLoS One 7: e31039.

Cruts M, Theuns J, Van Broeckhoven C. 2012. Locus-specific mutation databases for neurodegenerative brain diseases. Hum Mutat 33: 1340–1344.

Danecek P, Auton A, Abecasis G, Albers CA, Banks E, DePristo MA, Handsaker RE, Lunter G, Marth GT, Sherry ST, et al. 2011. The variant call format and VCFtools. Bioinformatics 27: 2156–2158.

Dolzhenko E, Deshpande V, Schlesinger F, Krusche P, Petrovski R, Chen S, Emig-Agius D, Gross A, Narzisi G, Bowman B, et al. 2019. ExpansionHunter: a sequence-graph-based tool to analyze variation in short tandem repeat regions. Bioinformatics 35: 4754–4756.

Dolzhenko E, van Vugt JJFA, Shaw RJ, Bekritsky MA, van Blitterswijk M, Narzisi G, Ajay SS, Rajan V, Lajoie BR, Johnson NH, et al. 2017. Detection of long repeat expansions from PCR-free whole-genome sequence data. Genome Res 27: 1895– 1903.

Dua T, Seeher KM, Sivananthan S, Chowdhary N, Pot AM, Saxena S. 2017. [FTS5–03– 01]: WORLD HEALTH ORGANIZATION’s GLOBAL ACTION PLAN ON THE PUBLIC HEALTH RESPONSE TO DEMENTIA 2017–2025. Alzheimer’s & Dementia 13. http://dx.doi.org/10.1016/j.jalz.2017.07.758.

Farrer LA, Adrienne Cupples L, Haines JL, Hyman B, Kukull WA, Mayeux R, Myers RH, Pericak-Vance MA, Risch N, van Duijn CM. 1997. Effects of Age, Sex, and Ethnicity on the Association Between Apolipoprotein E Genotype and Alzheimer Disease: A Meta-analysis. JAMA 278: 1349–1356.

Goldman JS, Farmer JM, Wood EM, Johnson JK, Boxer A, Neuhaus J, Lomen-Hoerth C, Wilhelmsen KC, Lee VM-Y, Grossman M, et al. 2005. Comparison of family histories in FTLD subtypes and related tauopathies. Neurology 65: 1817–1819.

Holstege H, Hulsman M, Charbonnier C, Grenier-Boley B, Quenez O, Grozeva D, van Rooij JGJ, Sims R, Ahmad S, Amin N, et al. 2021. Exome sequencing identifies rare damaging variants in the ATB8B4 and ABCA1 genes as novel risk factors for Alzheimer’s disease. Alzheimers Dement 17 Suppl 3: e055982.

Holstege H, van der Lee SJ, Hulsman M, Wong TH, van Rooij JG, Weiss M, Louwersheimer E, Wolters FJ, Amin N, Uitterlinden AG, et al. 2017. Characterization of pathogenic SORL1 genetic variants for association with Alzheimer’s disease: a clinical interpretation strategy. Eur J Hum Genet 25: 973– 981.

Holt JM, Wilk B, Birch CL, Brown DM, Gajapathy M, Moss AC, Sosonkina N, Wilk MA, Anderson JA, Harris JM, et al. 2019. VarSight: prioritizing clinically reported variants with binary classification algorithms. BMC Bioinformatics 20: 496.

Hsu S, Gordon BA, Hornbeck R, Norton JB, Levitch D, Louden A, Ziegemeier E, Laforce R Jr, Chhatwal J, Day GS, et al. 2018. Discovery and validation of autosomal dominant Alzheimer’s disease mutations. Alzheimers Res Ther 10: 67.

Hsu S, Pimenova AA, Hayes K, Villa JA, Rosene MJ, Jere M, Goate AM, Karch CM. 2020. Systematic validation of variants of unknown significance in APP, PSEN1 and PSEN2. Neurobiol Dis 139: 104817.

Huq AJ, Thompson B, Bennett MF, Bournazos A, Bommireddipalli S, Gorelik A, Schultz J, Sexton A, Purvis R, West K, et al. 2022. Clinical impact of whole-genome sequencing in patients with early-onset dementia. J Neurol Neurosurg Psychiatry. http://dx.doi.org/10.1136/jnnp-2021-328146.

Jian X, Boerwinkle E, Liu X. 2014. In silico prediction of splice-altering single nucleotide variants in the human genome. Nucleic Acids Res 42: 13534–13544.

Kapplinger JD, Tester DJ, Alders M, Benito B, Berthet M, Brugada J, Brugada P, Fressart V, Guerchicoff A, Harris-Kerr C, et al. 2010. An international compendium of mutations in the SCN5A-encoded cardiac sodium channel in patients referred for Brugada syndrome genetic testing. Heart Rhythm 7: 33–46.

Karczewski KJ, Francioli LC, Tiao G, Cummings BB, Alföldi J, Wang Q, Collins RL, Laricchia KM, Ganna A, Birnbaum DP, et al. 2020. The mutational constraint spectrum quantified from variation in 141,456 humans. Nature 581: 434–443.

Kelleher RJ 3rd, Shen J. 2017. Presenilin-1 mutations and Alzheimer’s disease. Proc Natl Acad Sci U S A 114: 629–631.

Kendig KI, Baheti S, Bockol MA, Drucker TM, Hart SN, Heldenbrand JR, Hernaez M, Hudson ME, Kalmbach MT, Klee EW, et al. 2019. Sentieon DNASeq Variant Calling Workflow Demonstrates Strong Computational Performance and Accuracy. Front Genet 10: 736.

Kim S, Jhong J-H, Lee J, Koo J-Y. 2017. Erratum to: Meta-analytic support vector machine for integrating multiple omics data. BioData Min 10: 8.

Kim S, Scheffler K, Halpern AL, Bekritsky MA, Noh E, Källberg M, Chen X, Kim Y, Beyter D, Krusche P, et al. 2018. Strelka2: fast and accurate calling of germline and somatic variants. Nat Methods 15: 591–594.

Kircher M, Witten DM, Jain P, O’Roak BJ, Cooper GM, Shendure J. 2014. A general framework for estimating the relative pathogenicity of human genetic variants. Nat Genet 46: 310–315.

Kunkle BW, Schmidt M, Klein H-U, Naj AC, Hamilton-Nelson KL, Larson EB, Evans DA, De Jager PL, Crane PK, Buxbaum JD, et al. 2021. Novel Alzheimer Disease Risk Loci and Pathways in African American Individuals Using the African Genome Resources Panel: A Meta-analysis. JAMA Neurol 78: 102–113.

Lambert JC, Ibrahim-Verbaas CA, Harold D, Naj AC, Sims R, Bellenguez C, DeStafano AL, Bis JC, Beecham GW, Grenier-Boley B, et al. 2013. Meta-analysis of 74,046 individuals identifies 11 new susceptibility loci for Alzheimer’s disease. Nat Genet 45: 1452–1458.

Lek M, Karczewski KJ, Minikel EV, Samocha KE, Banks E, Fennell T, O’Donnell-Luria AH, Ware JS, Hill AJ, Cummings BB, et al. 2016. Analysis of protein-coding genetic variation in 60,706 humans. Nature 536: 285–291.

Liao X, Cai F, Sun Z, Zhang Y, Wang J, Jiao B, Guo J, Li J, Liu X, Guo L, et al. 2020. Identification of Alzheimer’s disease-associated rare coding variants in the ECE2 gene. JCI Insight 5. http://dx.doi.org/10.1172/jci.insight.135119.

Li H. 2013. Aligning sequence reads, clone sequences and assembly contigs with BWA-MEM. arXiv [q-bioGN]. http://arxiv.org/abs/1303.3997.

Luckett ES, Abakkouy Y, Reinartz M, Adamczuk K, Schaeverbeke J, Verstockt S, De Meyer S, Van Laere K, Dupont P, Cleynen I, et al. 2022. Association of Alzheimer’s disease polygenic risk scores with amyloid accumulation in cognitively intact older adults. Alzheimers Res Ther 14: 138.

Manichaikul A, Mychaleckyj JC, Rich SS, Daly K, Sale M, Chen W-M. 2010. Robust relationship inference in genome-wide association studies. Bioinformatics 26: 2867– 2873.

Mukherjee O, Wang J, Gitcho M, Chakraverty S, Taylor-Reinwald L, Shears S, Kauwe JSK, Norton J, Levitch D, Bigio EH, et al. 2008. Molecular characterization of novel progranulin (GRN) mutations in frontotemporal dementia. Hum Mutat 29: 512–521.

Ng PC, Henikoff S. 2003. SIFT: Predicting amino acid changes that affect protein function. Nucleic Acids Res 31: 3812–3814.

NHLBI UM. 2018. The NHLBI Trans-Omics for Precision Medicine (TOPMed) Whole Genome Sequencing Program. BRAVO variant browser.

Pashaei E, Ozen M, Aydin N. 2016. Splice sites prediction of human genome using AdaBoost. In 2016 IEEE-EMBS International Conference on Biomedical and Health Informatics (BHI), pp. 300–303.

Piazza M, Prior TW, Khalsa PS, Appleby B. 2020. A case report of genetic prion disease with two different PRNP variants. Mol Genet Genomic Med 8: e1134.

Rademakers R, Dermaut B, Peeters K, Cruts M, Heutink P, Goate A, Van Broeckhoven C. 2003. Tau (MAPT) mutation Arg406Trp presenting clinically with Alzheimer disease does not share a common founder in Western Europe. Hum Mutat 22: 409–411.

Raghavan NS, Brickman AM, Andrews H, Manly JJ, Schupf N, Lantigua R, Wolock CJ, Kamalakaran S, Petrovski S, Tosto G, et al. 2018. Whole-exome sequencing in 20,197 persons for rare variants in Alzheimer’s disease. Annals of Clinical and Translational Neurology 5: 832–842. http://dx.doi.org/10.1002/acn3.582.

Rausch T, Zichner T, Schlattl A, Stütz AM, Benes V, Korbel JO. 2012. DELLY: structural variant discovery by integrated paired-end and split-read analysis. Bioinformatics 28: i333–i339.

Reed LA, Grabowski TJ, Schmidt ML, Morris JC, Goate A, Solodkin A, Van Hoesen GW, Schelper RL, Talbot CJ, Wragg MA, et al. 1997. Autosomal dominant dementia with widespread neurofibrillary tangles. Ann Neurol 42: 564–572.

Reitz C, Jun G, Naj A, Rajbhandary R, Vardarajan BN, Wang L-S, Valladares O, Lin C-F, Larson EB, Graff-Radford NR, et al. 2013. Variants in the ATP-binding cassette transporter (ABCA7), apolipoprotein E ϵ4,and the risk of late-onset Alzheimer disease in African Americans. JAMA 309: 1483–1492.

Richards S, Aziz N, Bale S, Bick D, Das S, Gastier-Foster J, Grody WW, Hegde M, Lyon E, Spector E, et al. 2015. Standards and guidelines for the interpretation of sequence variants: a joint consensus recommendation of the American College of Medical Genetics and Genomics and the Association for Molecular Pathology. Genet Med 17: 405–424.

Rosini F, Rufa A, Monti L, Tirelli L, Federico A. 2014. Adult-onset phenylketonuria revealed by acute reversible dementia, prosopagnosia and parkinsonism. J Neurol 261: 2446–2448.

Shankaran SS, Capell A, Hruscha AT, Fellerer K, Neumann M, Schmid B, Haass C. 2008. Missense mutations in the progranulin gene linked to frontotemporal lobar degeneration with ubiquitin-immunoreactive inclusions reduce progranulin production and secretion. J Biol Chem 283: 1744–1753.

Sinha N, Reagh ZM, Tustison NJ, Berg CN, Shaw A, Myers CE, Hill D, Yassa MA, Gluck MA. 2019. ABCA7 risk variant in healthy older African Americans is associated with a functionally isolated entorhinal cortex mediating deficient generalization of prior discrimination training. Hippocampus 29: 527–538.

Stoychev KR, Stoimenova-Popova M, Chumpalova P, Ilieva L, Swamad M, Kamburova-Martinova Z. 2019. A Clinical Case of Patient Carrying Rare Pathological PSEN1 Gene Mutation (L424V) Demonstrates the Phenotypic Heterogenity of Early Onset Familial AD. Front Psychiatry 10: 857.

Tufekcioglu Z, Cakar A, Bilgic B, Hanagasi H, Gurvit H, Emre M. 2016. Adult-onset phenylketonuria with rapidly progressive dementia and parkinsonism. Neurocase 22: 273–275.

van Spronsen FJ, Blau N, Harding C, Burlina A, Longo N, Bosch AM. 2021. Phenylketonuria. Nat Rev Dis Primers 7: 36.

Wickham H. 2016. ggplot2: Elegant Graphics for Data Analysis. Springer International Publishing.

Xi R, Lee S, Xia Y, Kim T-M, Park PJ. 2016. Copy number analysis of whole-genome data using BIC-seq2 and its application to detection of cancer susceptibility variants. Nucleic Acids Res 44: 6274–6286.

Zhu M, Need AC, Han Y, Ge D, Maia JM, Zhu Q, Heinzen EL, Cirulli ET, Pelak K, He M, et al. 2012. Using ERDS to infer copy-number variants in high-coverage genomes. Am J Hum Genet 91: 408–421.

2020. 2020 Alzheimer’s disease facts and figures. Alzheimers Dement. http://dx.doi.org/10.1002/alz.12068.

