## Supplemental Figure 1 - 5 for "Contributions of rare and common variation to early-onset and atypical dementia risk"

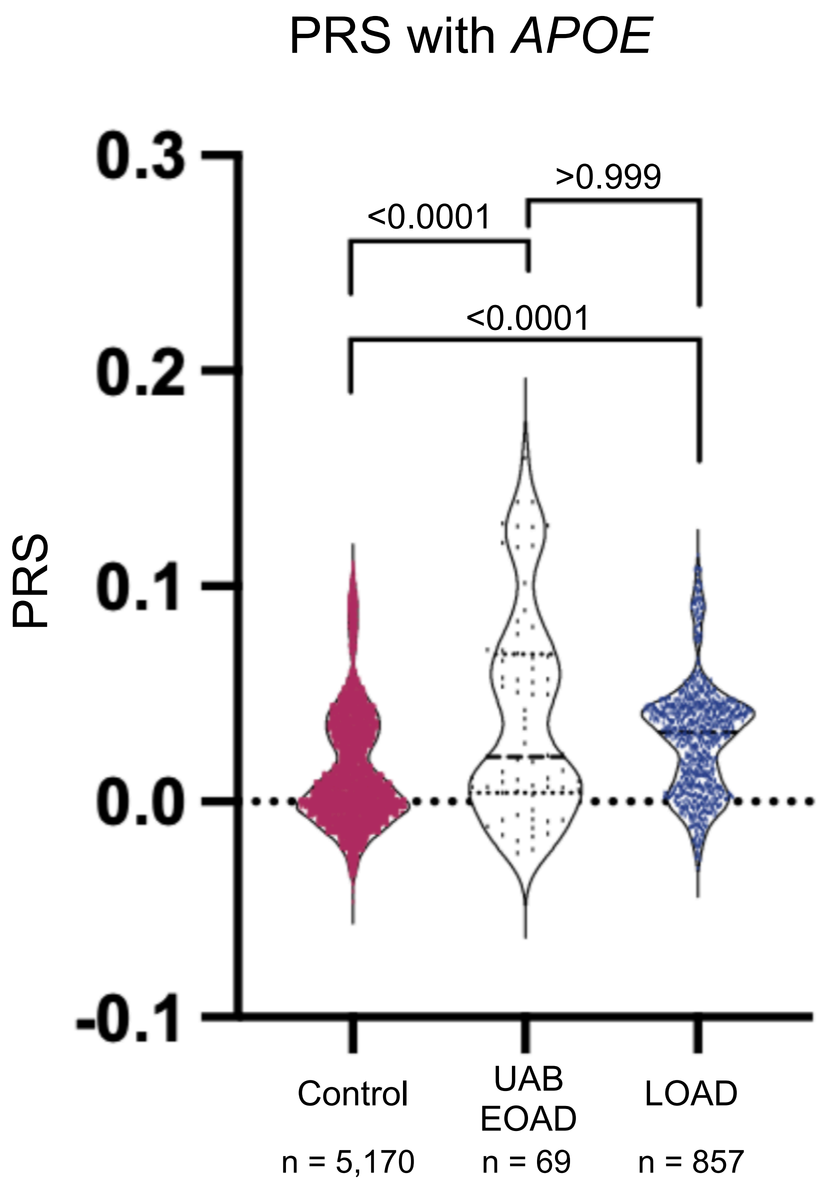


**Supplemental Figure 1.** Violin plot comparing polygenic risk scores calculated with *APOE* between the UAB early-onset Alzheimer's (UAB EOAD) cohort, late-onset Alzheimer's (LOAD) cohort, and the control (CTL) cohort. All individuals plotted were Caucasian, Non-Hispanic. Non-parametric ANOVA with Kruskal-Wallis test was used to calculate adjusted p-values.

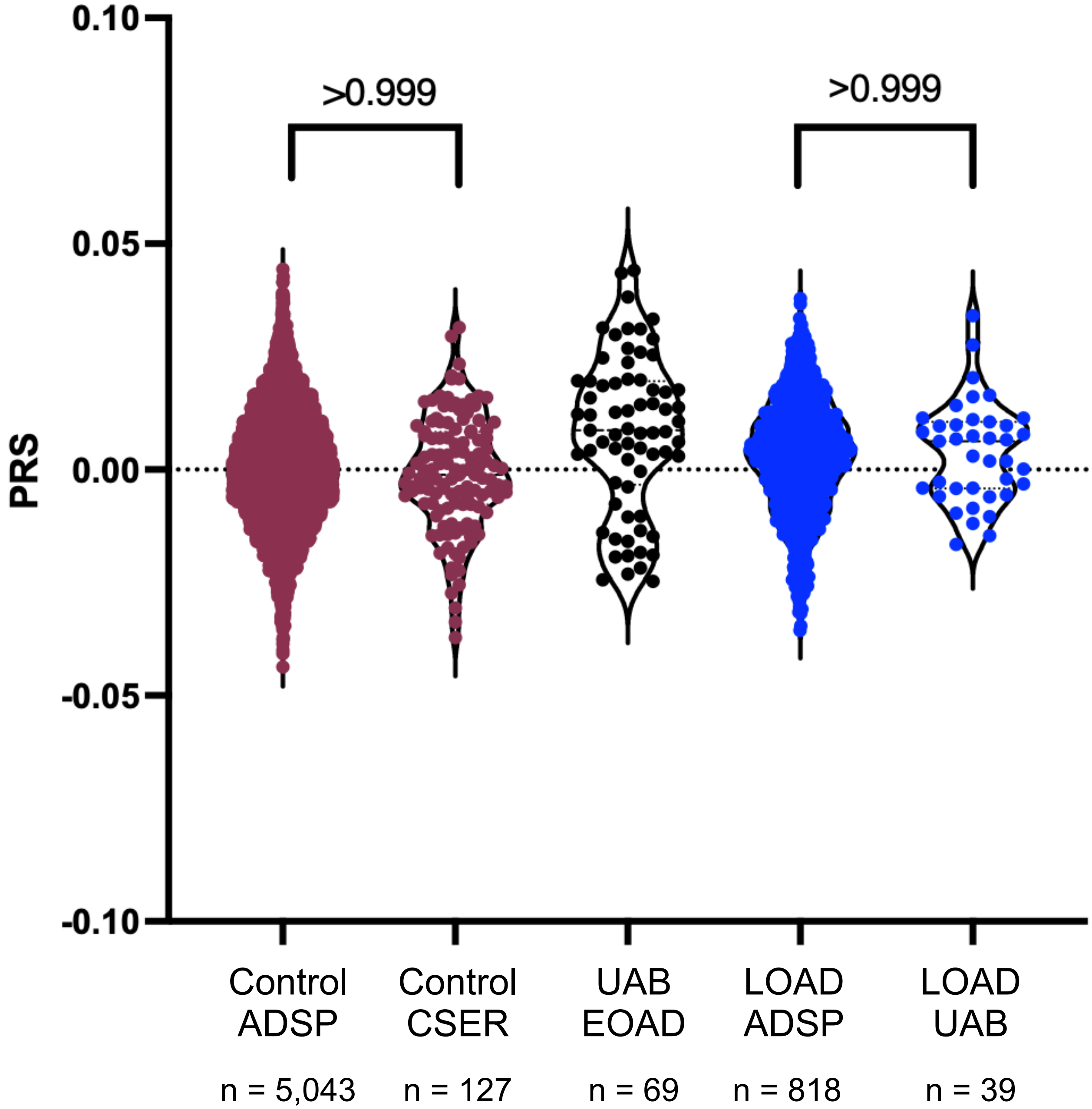


**Supplemental Figure 2.** Violin plot comparing polygenic risk scores between the UAB early-onset Alzheimer's (UAB EOAD) cohort, ADSP late-onset Alzheimer's (LOAD ADSP) and UAB late-onset dementia (LOAD UAB) cohorts, and the ADSP control (Control ADSP) and CSER control (Control CSER) cohorts. Y-axis represents polygenic risk score and x-axis is the respective cohort. This PRS excludes *APOE* status. All individuals plotted were Caucasian, Non-Hispanic. Non-parametric ANOVA with Kruskal-Wallis test was used to calculate adjusted p-values.

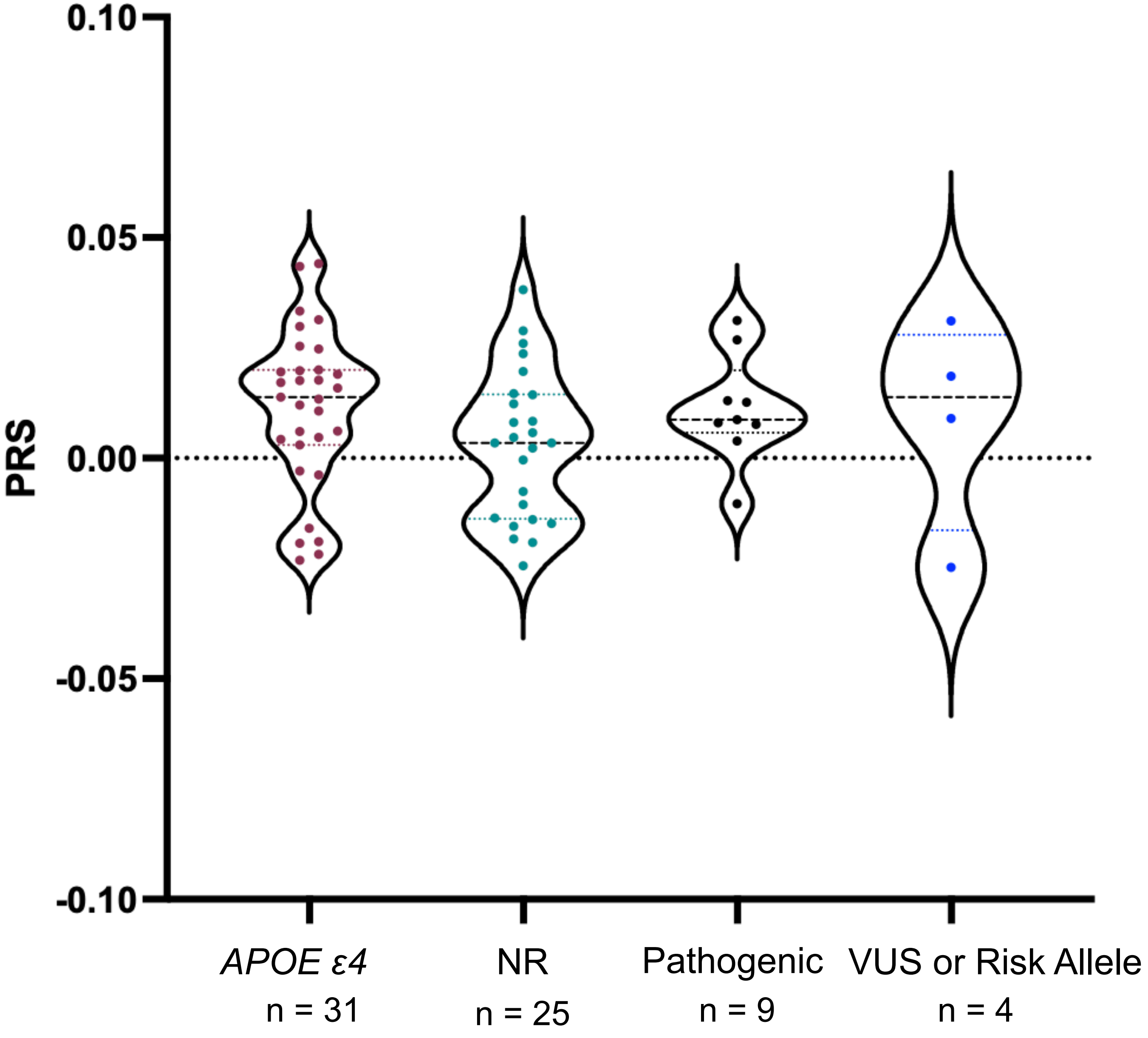


**Supplemental Figure 3.** Boxplot comparing PRS of UAB EOAD samples with a *APOE ε4* heterozygous or *APOE ε4* homozygous finding. NR = no returnable finding. VUS = variant of uncertain significance. This PRS excludes *APOE* status. All individuals plotted were Caucasian, Non-Hispanic. The p-value of the non-parametric ANOVA was 0.3323.

A

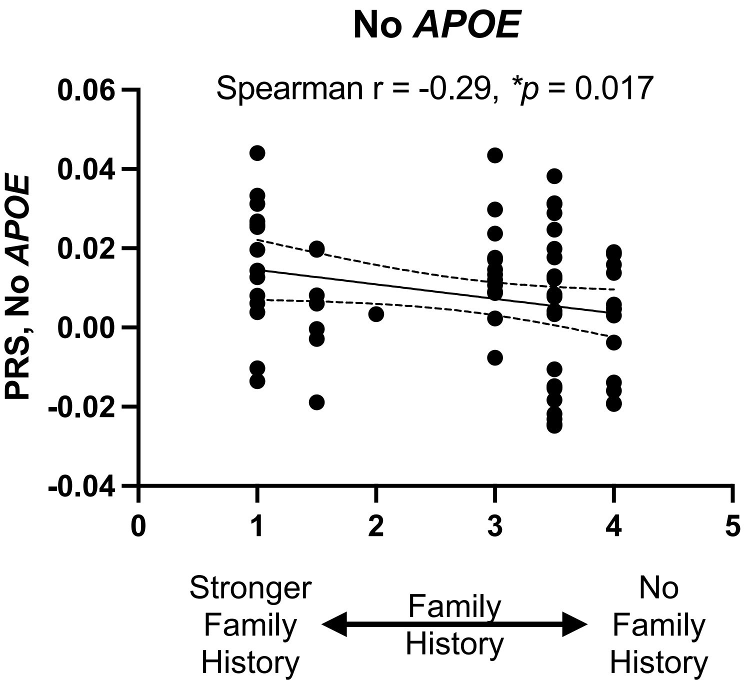


B

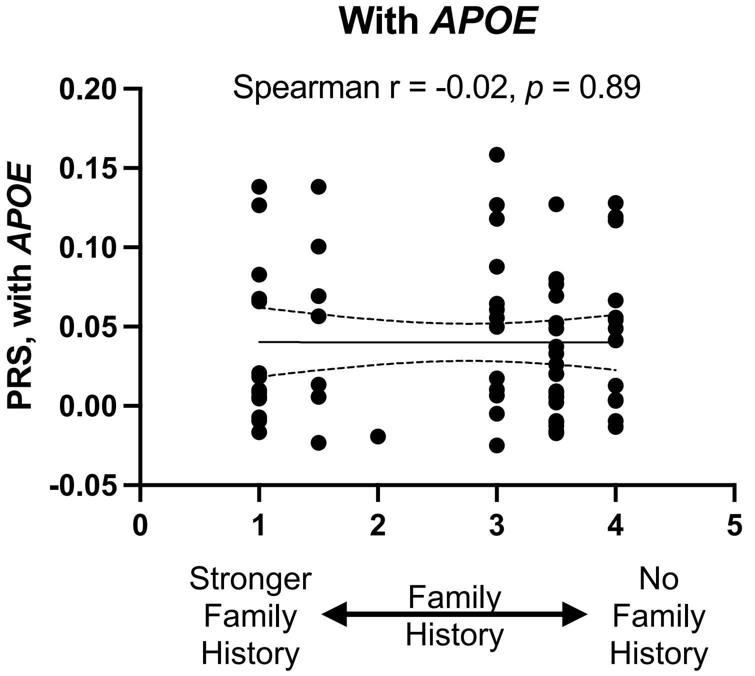


**Supplemental Figure 4.** Correlation plots comparing UAB EOAD family history scores and PRS (A) excluding and (B) including *APOE*. Y-axis represents polygenic risk score and x-axis is the respective family history score. Family history score = 1 is the strongest and 4 = no family history. All individuals plotted were Caucasian, Non-Hispanic. Lines represent +/- 95% confidence interval. Spearman's rank correlation test was used to calculate p-values.

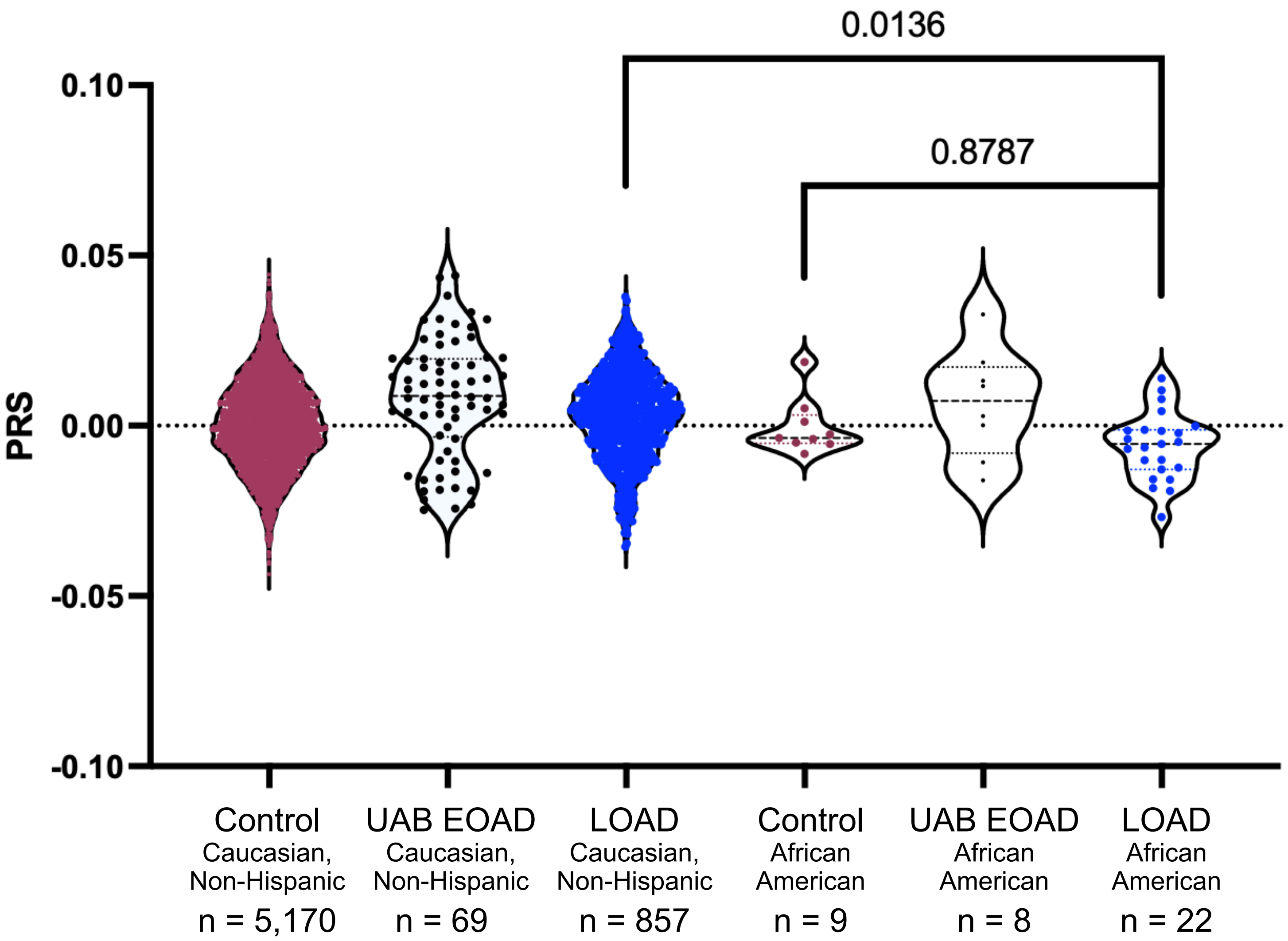


**Supplemental Figure 5.** Boxplot comparing PRS of control and Alzheimer's cohorts. Y-axis represents polygenic risk score and x-axis is the respective cohort. This PRS excludes *APOE* status. Non-parametric ANOVA with Kruskal-Wallis test was used to calculate adjusted p-values.
