## Supplemental Table 1 for "Contributions of rare and common variation to early-onset and atypical dementia risk"

| Clinical Assessment Before Testing | Decade of Onset | Family History Score | Ethnicity | Finding |
|---|---|---|---|---|
| FTD | 60's | 1 | African American, Non-Hispanic | See Cochran et al. |
| Unclear | 40's | 1 | Caucasian, Non-Hispanic | See Cochran et al. |
| EOAD | 60s | 1 | Caucasian, Non-Hispanic | See Cochran et al. |
| EOAD | 40's | 3.5 | Caucasian, Non-Hispanic | See Cochran et al. |
| EOAD vs FTD | 50's | 3.5 | Caucasian, Non-Hispanic | See Cochran et al. |
| FTD | 50's | 3 | Caucasian, Non-Hispanic | See Cochran et al. |
| FTD | 60's | 3.5 | Caucasian, Non-Hispanic | See Cochran et al. |
| EOAD v. FTD | 50s | 1 | Caucasian, Non-Hispanic | See Cochran et al. |
| FTD | 50's | 1.5 | Caucasian, Non-Hispanic | See Cochran et al. |
| EOAD v. FTD | 50's | 1 | Caucasian, Non-Hispanic | See Cochran et al. |
| EOAD | 40's | 3.5 | Caucasian, Non-Hispanic | See Cochran et al. |
| CBD | 50's | 3.5 | Caucasian, Non-Hispanic | See Cochran et al. |
| EOAD v. FTD | 40's | 4 | African American, Non-Hispanic | See Cochran et al. |
| EOAD | 40's | 4 | Caucasian, Non-Hispanic | See Cochran et al. |
| EOAD | 50's | 4 | Caucasian, Non-Hispanic | See Cochran et al. |
| EOAD | 40's | 3.5 | Caucasian, Non-Hispanic | See Cochran et al. |
| FTD | 40's | 3 | Caucasian, Non-Hispanic | See Cochran et al. |
| PPA | 50's | 4 | Caucasian, Non-Hispanic | See Cochran et al. |
| Dementia | 50's | 3 | Caucasian, Non-Hispanic | See Cochran et al. |
| EOAD | 40's | 3 | Caucasian, Non-Hispanic | See Cochran et al. |

| | | | | |
|---|---|---|---|---|
| FTD | 50's | 3 | Caucasian, Non-Hispanic | See Cochran et al. |
| Likely EOAD | 60's | 3.5 | Caucasian, Non-Hispanic | See Cochran et al. |
| Likely EOAD | 50's | 3 | Caucasian, Non-Hispanic | See Cochran et al. |
| EOAD | 50's | 3.5 | African American, Non-Hispanic | See Cochran et al. |
| EOAD | 40's | 1 | Caucasian, Non-Hispanic | See Cochran et al. |
| EOAD | 40's | 1 | Caucasian, Non-Hispanic | See Cochran et al. |
| EOAD | 50's | 1 | Caucasian, Non-Hispanic | See Cochran et al. |
| FTD | 40's | 3.5 | Caucasian, Non-Hispanic | See Cochran et al. |
| EOAD | 50's | 3.5 | Caucasian, Non-Hispanic | See Cochran et al. |
| CAA v. Leukodystrophy | 70's | 1.5 | Caucasian, Non-Hispanic | See Cochran et al. |
| FTD v. ALS | 50's | 1.5 | Caucasian, Non-Hispanic | See Cochran et al. |
| EOAD | 50's | 3.5 | African American, Non-Hispanic | See Cochran et al. |
| FTD | 60's | 3 | Caucasian, Non-Hispanic | *APOE ε4* Heterozygous |
| EOAD | 50s | 3 | Caucasian, Non-Hispanic | VUS |
| EOAD v. FTD | 40's | 3 | Caucasian, Non-Hispanic | No Returnables |
| FTD | 60s | 3.5 | Caucasian, Non-Hispanic | No Returnables |
| EOAD | 50's | 4 | Caucasian, Non-Hispanic | No Returnables |
| EOAD | 60's | 3.5 | Caucasian, Non-Hispanic | Risk |
| EOAD | 50's | 3 | Caucasian, Non-Hispanic | *APOE ε4* Heterozygous |
| MSA | 50's | 3.5 | Caucasian, Non-Hispanic | No Returnables |
| AD | 60's | 3.5 | Caucasian, Non-Hispanic | *APOE ε4* Heterozygous |
| Dementia | 60's | 4 | African American, Non-Hispanic | VUS |
| FTD | 50's | 3.5 | Caucasian, Non-Hispanic | No Returnables |

| | | | | |
|---|---|---|---|---|
| EOAD | 50's | 3 | Caucasian, Non-Hispanic | No Returnables |
| Leukodystrophy | 50's | 4 | Caucasian, Non-Hispanic | *APOE ε4* Heterozygous |
| PCA | 60's | 3.5 | Caucasian, Non-Hispanic | No Returnables |
| MS v. CADASIL | 40s | 4 | African American, Non-Hispanic | No Returnables |
| MCI | 50's | 1 | Caucasian, Non-Hispanic | Pathogenic |
| MCI | 50's | 3 | Caucasian, Non-Hispanic | *APOE ε4* Homozygous |
| Dementia | 40's | 4 | Caucasian, Non-Hispanic | *APOE ε4* Heterozygous |
| EOAD | 50's | 3.5 | Caucasian, Non-Hispanic | Pathogenic |
| CBS | 50's | 3 | Caucasian, Non-Hispanic | No Returnables |
| EOAD v. FTD | 50's | 3 | Caucasian, Non-Hispanic | Pathogenic |
| EOAD | 60's | 1.5 | Caucasian, Non-Hispanic | *APOE ε4* Homozygous |
| Dementia | 70's | 3 | Caucasian, Non-Hispanic | No Returnables |
| EOAD | 50's | 3.5 | Caucasian, Non-Hispanic | No Returnables |
| FTD | 40's | 3 | Caucasian, Non-Hispanic | No Returnables |
| Dementia | 60's | 1 | Caucasian, Non-Hispanic | No Returnables |
| FTD | 60's | 1 | Caucasian, Non-Hispanic | *APOE ε4* Heterozygous |
| EOAD | 60's | 4 | Caucasian, Non-Hispanic | *APOE ε4* Heterozygous |
| Dementia | 80s | 3.5 | Caucasian, Non-Hispanic | *APOE ε4* Heterozygous |
| Dementia | 50's | 3.5 | Caucasian, Non-Hispanic | *APOE ε4* Heterozygous |
| MCI | 50's | 1.5 | Caucasian, Non-Hispanic | No Returnables |
| Unclear | 40's | 4 | Caucasian, Non-Hispanic | No Returnables |
| Dementia | 60's | 4 | Caucasian, Non-Hispanic | *APOE ε4* Heterozygous |
| Dementia | 50's | 1.5 | Caucasian, Non-Hispanic | *APOE ε4* Heterozygous |

| | | | | |
|---|---|---|---|---|
| Non-dementia cognitive impairment | 50's | 3.5 | Caucasian, Non-Hispanic | No Returnables |
| EOAD | 40's | 3 | Caucasian, Non-Hispanic | VUS |
| Dementia | 30s | 2 | Caucasian, Non-Hispanic | No Returnables |
| Cognitive Impairment | 60's | 1.5 | Caucasian, Non-Hispanic | *APOE ε4* Homozygous |
| Logopenix PPA | 60's | 4 | Caucasian, Non-Hispanic | No Returnables |
| Dementia | 40s | 3.5 | Caucasian, Non-Hispanic | Pathogenic |
| Dementia | 60's | 1 | Caucasian, Non-Hispanic | No Returnables |
| EOAD | 50's | 4 | Caucasian, Non-Hispanic | VUS |
| MCI | 60's | 1 | Caucasian, Non-Hispanic | Pathogenic |
| EOAD v. FTD | 60's | 3.5 | Caucasian, Non-Hispanic | No Returnables |
| White Matter Disease | 50's | 3 | Caucasian, Non-Hispanic | *APOE ε4* Heterozygous |
| MCI | 50's | 1.5 | African American, Non-Hispanic | *APOE ε4* Heterozygous + Risk |
| EOAD | 50's | 3.5 | Caucasian, Non-Hispanic | No Returnables |
| EOAD | 50s | 4 | Caucasian, Non-Hispanic | *APOE ε4* Homozygous |
| EOAD | 40's | 3 | Caucasian, Non-Hispanic | *APOE ε4* Homozygous |
| EOAD | 50's | 3.5 | African American, Non-Hispanic | No Returnables |
| MCI | 60's | 3.5 | Caucasian, Non-Hispanic | *APOE ε4* Heterozygous |
| MCI | 60's | 3 | Caucasian, Non-Hispanic | No Returnables |
| EOAD | 60's | 3.5 | Caucasian, Non-Hispanic | No Returnables |
| EOAD | 50's | 1 | African American, Non-Hispanic | *APOE ε4* Heterozygous + Risk |
| MCI | 60's | 1.5 | Caucasian, Non-Hispanic | No Returnables |
| Early-onset dementia | 50s | 3.5 | Caucasian, Non-Hispanic | No Returnables |
| Early-onset dementia | 50's | 1.5 | African American, Non-Hispanic | No Returnables |

| | | | | |
|---|---|---|---|---|
| MCI | 70's | 1 | Caucasian, Non-Hispanic | *APOE ε4* Heterozygous + Risk |
| MCI | 70's | 1 | Caucasian, Non-Hispanic | *APOE ε4* Homozygous |
| EOAD v. FTD | 40's | 3.5 | Caucasian, Non-Hispanic | No Returnables |
| EOAD | 50's | 1 | Caucasian, Non-Hispanic | *APOE ε4* Heterozygous |
| Mild Dementia likely due to FTLD | 70's | 3 | Caucasian, Non-Hispanic | No Returnables |
| MCI | 60's | 1.5 | Caucasian, Non-Hispanic | *APOE ε4* Homozygous |
| EOAD | 60's | 4 | Caucasian, Non-Hispanic | *APOE ε4* Homozygous |
| MCI | 50's | 3.5 | Caucasian, Non-Hispanic | No Returnables |
| Progressive spastic dysarthria | 60's | 4 | Caucasian, Non-Hispanic | No Returnables |
| Dementia | 50's | 3.5 | Caucasian, Non-Hispanic | Risk |
| EOAD | 40's | 3 | Caucasian, Non-Hispanic | *APOE* ε4 Heterozygous + Risk |

Modified Goldman Score: (1) At least three people in two generations affected with EOAD, Frontotemporal dementia (FTD), ALS, corticobasal degeneration (CBD), Parkinson's disease (PD), or progressive supranuclear palsy (PSP) with one person being a first-degree relative of the other two, Criteria matching (1) but with late-onset Alzheimer disease (LOAD) instead of EOAD, (2) At least two relatives with dementia, FTD, ALS, CBD, PD, or PSP and criteria for autosomal dominant inheritance were not met, (3) A single affected first or second degree family member with early-onset dementia or at least two with FTD, ALS, CBD, PD, mild cognitive impairment (MCI), or PSP, (3.5) A single affected first or second degree family member with late-onset dementia, FTD, ALS, CBD, PD, MCI, or PSP, (4) non contributory family history or unknown family history.
