## Supplemental Table 2 for "Contributions of rare and common variation to early-onset and atypical dementia risk"

| ID | PRS | PRS with *APOE* | Family History Score | Ethnicity |
|---|---|---|---|---|
| SL250970 | 0.0333065 | 0.138135 | 1 | Caucasian, Non-Hispanic |
| SL250972 | 0.0311821 | 0.0207881 | 1 | Caucasian, Non-Hispanic |
| SL254259 | 0.0313604 | 0.0800914 | 3.5 | Caucasian, Non-Hispanic |
| SL260175 | 0.0083537 | 0.00589673 | 3.5 | Caucasian, Non-Hispanic |
| SL264669 | 0.00611485 | 0.00458614 | 1 | Caucasian, Non-Hispanic |
| SL261825 | -0.0103155 | -0.0070919 | 1 | Caucasian, Non-Hispanic |
| SL269480 | -0.0105164 | 0.0489248 | 3.5 | Caucasian, Non-Hispanic |
| SL269481 | -0.0231081 | 0.0330434 | 3.5 | Caucasian, Non-Hispanic |
| SL270813 | 0.0185293 | 0.134528 | 4 | African American, Non-Hispanic |
| SL271949 | 0.0159059 | 0.0665396 | 4 | Caucasian, Non-Hispanic |
| SL280004 | 0.0138158 | 0.0543399 | 4 | Caucasian, Non-Hispanic |
| SL278685 | 0.0247405 | 0.0694488 | 3.5 | Caucasian, Non-Hispanic |
| SL299914 | -0.0139039 | -0.009559 | 4 | Caucasian, Non-Hispanic |
| SL332858 | 0.0176364 | 0.0645647 | 3 | Caucasian, Non-Hispanic |
| SL318809 | 0.0171551 | 0.126734 | 3 | Caucasian, Non-Hispanic |
| SL332859 | 0.0176798 | 0.127105 | 3.5 | Caucasian, Non-Hispanic |
| SL338611 | 0.0434726 | 0.0878778 | 3 | Caucasian, Non-Hispanic |
| SL340577 | 9.98E-05 | 0.0525084 | 3.5 | African American, Non-Hispanic |
| SL340579 | 0.0268173 | 0.0184369 | 1 | Caucasian, Non-Hispanic |
| SL338612 | 0.00804379 | 0.00580941 | 1 | Caucasian, Non-Hispanic |
| SL346298 | -0.0243806 | -0.0172099 | 3.5 | Caucasian, Non-Hispanic |
| SL359223 | 0.0326719 | 0.144251 | 3.5 | African American, Non-Hispanic |
| SL367837 | 0.0090353 | 0.00665759 | 3 | Caucasian, Non-Hispanic |
| SL368913 | -0.0135294 | -0.0093015 | 1 | Caucasian, Non-Hispanic |
| SL372007 | 0.00577506 | 0.00385004 | 4 | Caucasian, Non-Hispanic |
| SL378766 | 0.031124 | 0.0200083 | 3.5 | Caucasian, Non-Hispanic |
| SL375559 | 0.0120798 | 0.0607446 | 3 | Caucasian, Non-Hispanic |

| | | | | |
|---|---|---|---|---|
| SL389240 | 0.00606756 | 0.0566112 | 1.5 | Caucasian, Non-Hispanic |
| SL393580 | 0.00284256 | 0.00200651 | 4 | African American, Non-Hispanic |
| SL403046 | 0.023696 | 0.0174602 | 3 | Caucasian, Non-Hispanic |
| SL399032 | -0.0038122 | 0.0413508 | 4 | Caucasian, Non-Hispanic |
| SL407847 | 0.00388921 | -0.0165281 | 1 | Caucasian, Non-Hispanic |
| SL409500 | 0.0133924 | 0.117929 | 3 | Caucasian, Non-Hispanic |
| SL411990 | 0.0030056 | 0.0487838 | 4 | Caucasian, Non-Hispanic |
| SL409501 | 0.00769402 | 0.00543107 | 3.5 | Caucasian, Non-Hispanic |
| SL414239 | 0.00868406 | 0.055485 | 3 | Caucasian, Non-Hispanic |
| SL419860 | -0.0028926 | 0.100428 | 1.5 | Caucasian, Non-Hispanic |
| SL423440 | 0.0146096 | 0.0103126 | 3 | Caucasian, Non-Hispanic |
| SL419861 | 0.00341336 | 0.0021943 | 3.5 | Caucasian, Non-Hispanic |
| SL420859 | 0.025975 | 0.0676904 | 1 | Caucasian, Non-Hispanic |
| SL431896 | 0.019068 | 0.128084 | 4 | Caucasian, Non-Hispanic |
| SL446286 | -0.0217881 | 0.0374604 | 3.5 | Caucasian, Non-Hispanic |
| SL446287 | 0.0198614 | 0.0767636 | 3.5 | Caucasian, Non-Hispanic |
| SL446288 | -0.0003954 | -0.023302 | 1.5 | Caucasian, Non-Hispanic |
| SL447318 | -0.0190986 | -0.0131303 | 4 | Caucasian, Non-Hispanic |
| SL446289 | 0.00472065 | 0.0556852 | 4 | Caucasian, Non-Hispanic |
| SL450729 | 0.0199974 | 0.0692674 | 1.5 | Caucasian, Non-Hispanic |
| SL450265 | 0.00341484 | -0.019265 | 2 | Caucasian, Non-Hispanic |
| SL450730 | -0.0189114 | 0.138256 | 1.5 | Caucasian, Non-Hispanic |
| SL455090 | 0.00470161 | 0.00313441 | 4 | Caucasian, Non-Hispanic |
| SL463503 | 0.0129963 | 0.00917386 | 3.5 | Caucasian, Non-Hispanic |
| HALB3003530 | 0.0143336 | 0.00985436 | 1 | Caucasian, Non-Hispanic |
| HALB3004851 | 0.0185515 | 0.0127542 | 4 | Caucasian, Non-Hispanic |
| HALB3004850 | 0.0126843 | 0.00845617 | 1 | Caucasian, Non-Hispanic |
| HALB3002363 | -0.0183136 | -0.0125906 | 3.5 | Caucasian, Non-Hispanic |
| HALB3003295 | 0.0131676 | 0.0614924 | 1.5 | African American, Non-Hispanic |
| HALB3003262 | -0.0147982 | -0.0095132 | 3.5 | Caucasian, Non-Hispanic |
| HALB3003260 | -0.0159186 | 0.119296 | 4 | Caucasian, Non-Hispanic |

| | | | | |
|---|---|---|---|---|
| HALB3003258 | 0.0298254 | 0.158361 | 3 | Caucasian, Non-Hispanic |
| HALB3004409 | -0.0108189 | -0.007438 | 3.5 | African American, Non-Hispanic |
| HALB3004581 | 0.00422115 | 0.0523347 | 3.5 | Caucasian, Non-Hispanic |
| HALB3004582 | 0.00227368 | -0.0248585 | 3 | Caucasian, Non-Hispanic |
| HALB3005193 | 0.0122719 | 0.00818124 | 3.5 | Caucasian, Non-Hispanic |
| HALB3005426 | 0.0116371 | 0.0717025 | 1 | African American, Non-Hispanic |
| HALB3005194 | 0.00810661 | 0.00572231 | 1.5 | Caucasian, Non-Hispanic |
| HALB3010348 | 0.028915 | 0.0204106 | 3.5 | Caucasian, Non-Hispanic |
| HALB3009833 | -0.0160448 | -0.0366347 | 1.5 | African American, Non-Hispanic |
| HALB3011654 | 0.0195863 | 0.0659053 | 1 | Caucasian, Non-Hispanic |
| HALB3013405 | 0.0253773 | 0.126585 | 1 | Caucasian, Non-Hispanic |
| HALB3012099 | -0.0154113 | -0.0102742 | 3.5 | Caucasian, Non-Hispanic |
| HALB3015765 | 0.0440723 | 0.0827395 | 1 | Caucasian, Non-Hispanic |
| HALB3015595 | -0.0075837 | -0.0048753 | 3 | Caucasian, Non-Hispanic |
| HALB3015596 | 0.0196619 | 0.0135176 | 1.5 | Caucasian, Non-Hispanic |
| HALB3017454 | -0.0193005 | 0.117041 | 4 | Caucasian, Non-Hispanic |
| HALB3017455 | 0.0381732 | 0.0262441 | 3.5 | Caucasian, Non-Hispanic |
| HALB3019252 | -0.0247508 | -0.0159112 | 3.5 | Caucasian, Non-Hispanic |
| HALB3018069 | 0.0106806 | 0.0499623 | 3 | Caucasian, Non-Hispanic |
