## Supplemental ACMG Pathogenicity Evidence Details and Extended Acknowledgements for "Contributions of rare and common variation to early-onset and atypical dementia risk"

**SUPPLEMENTAL MATERIAL**

**ACMG Pathogenicity Evidence Details**

**GRN (NM_002087.3:c.26C>A, p.(Ala9Asp))**

- The specific missense *GRN* variant identified was not observed in a population database of healthy individuals, which is consistent with the expected frequency of a pathogenic variant in this gene. (ACMG criterion PM2)
- This specific variant has been reported in several different families with frontotemporal dementia (Chen-Plotkin et al. 2011; Shankaran et al. 2008; Mukherjee et al. 2008). It has been shown to segregate with disease in those families. (ACMG criterion PP1)
- Multiple lines of computational evidence support a deleterious effect of this variant. (ACMG criterion PP3)
- This variant has been found through functional studies to damage the gene or gene-product (Mukherjee et al. 2008; Shankaran et al. 2008) (ACMG criterion PS3)
- Same amino acid change as a previously established pathogenic variant. (ACMG criterion PS1)
- The presence of two strong criteria, two supporting criteria, and one moderate criterion result in the ACMG-recommended assertion of “pathogenic”.

**PSEN1 (NM_000021.4:c.236C>T, p.(Ala79Val))**

- p.Ala79Val has been previously reported as pathogenic (Cruchaga et al. 2012; Wallon et al. 2012). (ACMG criterion PS1)
- The c.236C>T variant has been shown to damage the gene or gene-product based on a well-established functional assay, amyloid beta ELISA. (ACMG criterion PS3)
- The Genome Aggregation Database (gnomAD) reports that the c.236C>T variant was observed in 5/143286 and 3/251430 alleles for genome and exome data, respectively; consistent with the expected frequency of a pathogenic variant in Alzheimer disease. (ACMG criterion PM2)
- The c.236C>T variant in gene PSEN1 co-segregates with Alzheimer disease in multiple affected family members related to the patient. (ACMG criterion PP1)
- Multiple lines of computational evidence support a deleterious effect of the p.Ala79Val variant on the gene or gene product. (ACMG criterion PP3)
- The presence of two strong criteria, two supporting criteria, and one moderate criterion result in the ACMG-recommended assertion of “pathogenic”.

**MAPT (NM_005910.5:c.1216C>T, p.(Arg406Trp))**

- The Genome Aggregation Database (gnomAD) reports that the c.236C>T variant was observed in 0/143286 and 4/251284 alleles for genome and exome data, respectively; consistent with the expected frequency of a pathogenic variant in Alzheimer’s disease. (ACMG criterion PM2)

- Predicted damaging by multiple computational methods (CADD (Kircher et al. 2014), PolyPhen-2 (Adzhubei et al. 2010), and SIFT (Ng and Henikoff 2003)) (ACMG criterion PP3).
- This variant has been found through functional studies to damage the gene or gene-product (Hasegawa et al. 1998; Hong et al. 1998; Krishnamurthy and Johnson 2004; 101 Zhang et al. 2004) (ACMG criterion PS3).
- Strong segregation with EOAD in multiple studies (Reed et al. 1997; Rademakers et al. 2003; 99 Cruts et al. 2012) (ACMG criterion PP1S).
- The presence of two strong criteria, one supporting criterion, and one moderate criterion result in the ACMG-recommended assertion of “pathogenic”.

**PAH (NM_000277.3:c.117C>G, p.(Phe39Leu))**

- The c.117C>G variant has been shown to damage the gene or gene-product based on a well-established functional assay. Binding assay with tetrahydrobiopterin (Erlandsen, 2004). (ACMG criterion PS3)
- The prevalence of the c.117C>G variant in affected individuals with Phenylketonuria is significantly increased compared to the prevalence in controls (O'Donnell, 2002). (ACMG criterion PS4)
- The Genome Aggregation Database (gnomAD) reports that the c.117C>G variant was observed in 13/143294 and 24/251412 alleles for genome and exome data, respectively; consistent with the expected frequency of a pathogenic variant in phenylketonuria. (ACMG criterion PM2)
- The c.117C>G variant was observed to be in trans with a pathogenic variant based on segregation analysis in the family. (ACMG criterion PM3)

- Multiple lines of computational evidence support a deleterious effect of the p.Phe39Leu variant on the gene or gene product. (ACMG criterion PP3)
- This patient's phenotype or family history are specifically associated with phenylketonuria where pathogenic variants in the PAH gene are the only known cause. (ACMG criterion PP4)
- The c.117C>G variant has been reported as pathogenic by a reputable source (ClinVar Variation ID 636); however, the primary data for this classification are not available for review. (ACMG criterion PP5)
- This collective evidence supports c.117C>G as a pathogenic variant for Phenylketonuria.

**PAH (NM_000277.3:c.194T>C, p.(Ile65Thr))**

- The c.194T>C variant has been shown to damage the gene or gene-product based on a well-established functional assay:binding assay with tetrahydrobiopterin (Erlandsen, 2004). (ACMG criterion PS3)
- The prevalence of the c.194T>C variant in affected individuals with Phenylketonuria is significantly increased compared to the prevalence in controls (O'Donnell, 2002). (ACMG criterion PS4)
- The Genome Aggregation Database (gnomAD) reports that the c.194T>C variant was observed in 45/143316 and 68/251202 alleles for genome and exome data, respectively; consistent with the expected frequency of a pathogenic variant in Phenylketonuria. (ACMG criterion PM2)
- The c.194T>C variant was observed to be in trans with a pathogenic variant based on segregation analysis in the family. Multiple lines of computational

evidence support a deleterious effect of the p.Ile65Thr variant on the gene or gene product. (ACMG criterion PM3)

- This patient's phenotype or family history are specifically associated with Phenylketonuria where pathogenic variants in the PAH gene are the only known cause. (ACMG criterion PP4)
- This collective evidence supports c.194T>C as a pathogenic variant for Phenylketonuria.

**SCN5A (NM_198056.2:c.673C>T, p.(Arg225Trp))**

- The Genome Aggregation Database (gnomAD) reports that the c.673C>T variant was observed in 12/152200 and 3/242066 alleles for genome and exome data, respectively; consistent with the expected frequency of a pathogenic variant in Brugada syndrome. (ACMG criterion PM2)
- Multiple lines of computational evidence support a deleterious effect of the p.Phe39Leu variant on the gene or gene product. (ACMG criterion PP3)
- Same amino acid change as a previously established pathogenic variant. (ACMG criterion PS1)
- Non-truncating, non-synonymous variant is located in a mutational hot spot and/or critical and well-established functional domain. (ACMG criterion PM1)
- This collective evidence supports c.673C>T as a pathogenic variant for Brugada syndrome.

### Extended Acknowledgements

#### ADSP:

*The Alzheimer's Disease Sequencing Project (ADSP) is comprised of two Alzheimer's Disease (AD) genetics consortia and three National Human Genome Research Institute (NHGRI) funded Large Scale Sequencing and Analysis Centers (LSAC). The two AD genetics consortia are the Alzheimer's Disease Genetics Consortium (ADGC) funded by NIA (U01 AG032984), and the Cohorts for Heart and Aging Research in Genomic Epidemiology (CHARGE) funded by NIA (R01 AG033193), the National Heart, Lung, and Blood Institute (NHLBI), other National Institute of Health (NIH) institutes and other foreign governmental and non-governmental organizations. The Discovery Phase analysis of sequence data is supported through UF1AG047133 (to Drs. Schellenberg, Farrer, Pericak-Vance, Mayeux, and Haines); U01AG049505 to Dr. Seshadri; U01AG049506 to Dr. Boerwinkle; U01AG049507 to Dr. Wijsman; and U01AG049508 to Dr. Goate and the Discovery Extension Phase analysis is supported through U01AG052411 to Dr. Goate, U01AG052410 to Dr. Pericak-Vance and U01 AG052409 to Drs. Seshadri and Fornage.*

*Sequencing for the Follow Up Study (FUS) is supported through U01AG057659 (to Drs. PericakVance, Mayeux, and Vardarajan) and U01AG062943 (to Drs. Pericak-Vance and Mayeux). Data generation and harmonization in the Follow-up Phase is supported by U54AG052427 (to Drs. Schellenberg and Wang). The FUS Phase analysis of sequence data is supported through U01AG058589 (to Drs. Destefano, Boerwinkle, De Jager, Fornage, Seshadri, and Wijsman), U01AG058654 (to Drs. Haines, Bush, Farrer, Martin, and Pericak-Vance), U01AG058635 (to Dr. Goate), RF1AG058066 (to Drs. Haines, Pericak-Vance, and Scott), RF1AG057519 (to Drs. Farrer and Jun), R01AG048927 (to Dr. Farrer), and RF1AG054074 (to Drs. Pericak-Vance and Beecham).*

*The ADGC cohorts include: Adult Changes in Thought (ACT) (U01 AG006781, U19 AG066567), the Alzheimer's Disease Research Centers (ADRC) (P30 AG062429, P30 AG066468, P30 AG062421, P30 AG066509, P30 AG066514, P30 AG066530, P30 AG066507, P30 AG066444, P30 AG066518, P30 AG066512, P30 AG066462, P30 AG072979, P30 AG072972, P30 AG072976, P30 AG072975, P30 AG072978, P30 AG072977, P30 AG066519, P30 AG062677, P30 AG079280, P30 AG062422, P30 AG066511, P30 AG072946, P30 AG062715, P30 AG072973, P30 AG066506, P30 AG066508, P30 AG066515, P30 AG072947, P30 AG072931, P30 AG066546, P20 AG068024, P20 AG068053, P20 AG068077, P20 AG068082, P30 AG072958, P30 AG072959), the Chicago Health and Aging Project (CHAP) (R01 AG11101, RC4 AG039085, K23 AG030944), Indiana Memory and Aging Study (IMAS) (R01*

*AG019771), Indianapolis Ibadan (R01 AG009956, P30 AG010133), the Memory and Aging Project (MAP) ( R01 AG17917), Mayo Clinic (MAYO) (R01 AG032990, U01 AG046139, R01 NS080820, RF1 AG051504, P50 AG016574), Mayo Parkinson's Disease controls (NS039764, NS071674, 5RC2HG005605), University of Miami (R01 AG027944, R01 AG028786, R01 AG019085, IIRG09133827, A2011048), the Multi-Institutional Research in Alzheimer's Genetic Epidemiology Study (MIRAGE) (R01 AG09029, R01 AG025259), the National Centralized Repository for Alzheimer's Disease and Related Dementias (NCRAD) (U24 AG021886), the National Institute on Aging Late Onset Alzheimer's Disease Family Study (NIA- LOAD) (U24 AG056270), the Religious Orders Study (ROS) (P30 AG10161, R01 AG15819), the Texas Alzheimer's Research and Care Consortium (TARCC) (funded by the Darrell K Royal Texas Alzheimer's Initiative), Vanderbilt University/Case Western Reserve University (VAN/CWRU) (R01 AG019757, R01 AG021547, R01 AG027944, R01 AG028786, P01 NS026630, and Alzheimer's Association), the Washington Heights-Inwood Columbia Aging Project (WHICAP) (RF1 AG054023), the University of Washington Families (VA Research Merit Grant, NIA: P50AG005136, R01AG041797, NINDS: R01NS069719), the Columbia University Hispanic Estudio Familiar de Influencia Genetica de Alzheimer (EFIGA) (RF1 AG015473), the University of Toronto (UT) (funded by Wellcome Trust, Medical Research Council, Canadian Institutes of Health Research), and Genetic Differences (GD) (R01 AG007584). The CHARGE cohorts are supported in part by National Heart, Lung, and Blood Institute (NHLBI) infrastructure grant HL105756 (Psaty), RC2HL102419 (Boerwinkle) and the neurology working group is supported by the National Institute on Aging (NIA) R01 grant AG033193.*

*The CHARGE cohorts participating in the ADSP include the following: Austrian Stroke Prevention Study (ASPS), ASPS-Family study, and the Prospective Dementia Registry-Austria (ASPS/PRODEM-Aus), the Atherosclerosis Risk in Communities (ARIC) Study, the Cardiovascular Health Study (CHS), the Erasmus Rucphen Family Study (ERF), the Framingham Heart Study (FHS), and the Rotterdam Study (RS). ASPS is funded by the Austrian Science Fond (FWF) grant number P20545-P05 and P13180 and the Medical University of Graz. The ASPS-Fam is funded by the Austrian Science Fund (FWF) project I904), the EU Joint Programme – Neurodegenerative Disease Research (JPND) in frame of the BRIDGET project (Austria, Ministry of Science) and the Medical University of Graz and the Steiermärkische Krankenanstalten Gesellschaft. PRODEM-Austria is supported by the Austrian Research Promotion agency (FFG) (Project No. 827462) and by the Austrian National Bank (Anniversary Fund, project 15435. ARIC research is carried out as a collaborative study supported by NHLBI contracts (HHSN268201100005C, HHSN268201100006C, HHSN268201100007C, HHSN268201100008C, HHSN268201100009C, HHSN268201100010C, HHSN268201100011C, and HHSN268201100012C).*

*Neurocognitive data in ARIC is collected by U01 2U01HL096812, 2U01HL096814, 2U01HL096899, 2U01HL096902, 2U01HL096917 from the NIH (NHLBI, NINDS, NIA and NIDCD), and with previous brain MRI examinations funded by R01-HL70825 from the NHLBI. CHS research was supported by contracts HHSN268201200036C, HHSN268200800007C, N01HC55222, N01HC85079, N01HC85080, N01HC85081, N01HC85082, N01HC85083, N01HC85086, and grants U01HL080295 and U01HL130114 from the NHLBI with additional contribution from the National Institute of Neurological Disorders and Stroke (NINDS). Additional support was provided by R01AG023629, R01AG15928, and R01AG20098 from the NIA. FHS research is supported by NHLBI contracts N01-HC-25195 and HHSN268201500001I. This study was also supported by additional grants from the NIA (R01s AG054076, AG049607 and AG033040 and NINDS (R01 NS017950). The ERF study as a part of EUROSPAN (European Special Populations Research Network) was supported by European Commission FP6 STRP grant number 018947 (LSHG-CT-2006-01947) and also received funding from the European Community's Seventh Framework Programme (FP7/2007-2013)/grant agreement HEALTH-F4- 2007-201413 by the European Commission under the programme "Quality of Life and Management of the Living Resources" of 5th Framework Programme (no. QLG2-CT-2002- 01254). High-throughput analysis of the ERF data was supported by a joint grant from the Netherlands Organization for Scientific Research and the Russian Foundation for Basic Research (NWO-RFBR 047.017.043). The Rotterdam Study is funded by Erasmus Medical Center and Erasmus University, Rotterdam, the Netherlands Organization for Health Research and Development (ZonMw), the Research Institute for Diseases in the Elderly (RIDE), the Ministry of Education, Culture and Science, the Ministry for Health, Welfare and Sports, the European Commission (DG XII), and the municipality of Rotterdam. Genetic data sets are also supported by the Netherlands Organization of Scientific Research NWO Investments (175.010.2005.011, 911-03-012), the Genetic Laboratory of the Department of Internal Medicine, Erasmus MC, the Research Institute for Diseases in the Elderly (014-93-015; RIDE2), and the Netherlands Genomics Initiative (NGI)/Netherlands Organization for Scientific Research (NWO) Netherlands Consortium for Healthy Aging (NCHA), project 050-060-810. All studies are grateful to their participants, faculty and staff. The content of these manuscripts is solely the responsibility of the authors and does not necessarily represent the official views of the National Institutes of Health or the U.S. Department of Health and Human Services.*

*The FUS cohorts include: the Alzheimer's Disease Research Centers (ADRC) (P30 AG062429, P30 AG066468, P30 AG062421, P30 AG066509, P30 AG066514, P30 AG066530, P30 AG066507, P30 AG066444, P30 AG066518, P30 AG066512, P30 AG066462, P30 AG072979, P30 AG072972, P30 AG072976, P30 AG072975, P30 AG072978, P30 AG072977, P30 AG066519, P30 AG062677, P30 AG079280, P30*

*AG062422, P30 AG066511, P30 AG072946, P30 AG062715, P30 AG072973, P30 AG066506, P30 AG066508, P30 AG066515, P30 AG072947, P30 AG072931, P30 AG066546, P20 AG068024, P20 AG068053, P20 AG068077, P20 AG068082, P30 AG072958, P30 AG072959), Alzheimer's Disease Neuroimaging Initiative (ADNI) (U19AG024904), Amish Protective Variant Study (RF1AG058066), Cache County Study (R01AG11380, R01AG031272, R01AG21136, RF1AG054052), Case Western Reserve University Brain Bank (CWRUBB) (P50AG008012), Case Western Reserve University Rapid Decline (CWRURD) (RF1AG058267, NU38CK000480), CubanAmerican Alzheimer's Disease Initiative (CuAADI) (3U01AG052410), Estudio Familiar de Influencia Genetica en Alzheimer (EFIGA) (5R37AG015473, RF1AG015473, R56AG051876), Genetic and Environmental Risk Factors for Alzheimer Disease Among African Americans Study (GenerAAtions) (2R01AG09029, R01AG025259, 2R01AG048927), Gwangju Alzheimer and Related Dementias Study (GARD) (U01AG062602), Hillblom Aging Network (2014-A-004-NET, R01AG032289, R01AG048234), Hussman Institute for Human Genomics Brain Bank (HIHGBB) (R01AG027944, Alzheimer's Association "Identification of Rare Variants in Alzheimer Disease"), Ibadan Study of Aging (IBADAN) (5R01AG009956), Longevity Genes Project (LGP) and LonGenity (R01AG042188, R01AG044829, R01AG046949, R01AG057909, R01AG061155, P30AG038072), Mexican Health and Aging Study (MHAS) (R01AG018016), Multi-Institutional Research in Alzheimer's Genetic Epidemiology (MIRAGE) (2R01AG09029, R01AG025259, 2R01AG048927), Northern Manhattan Study (NOMAS) (R01NS29993), Peru Alzheimer's Disease Initiative (PeADI) (RF1AG054074), Puerto Rican 1066 (PR1066) (Wellcome Trust (GR066133/GR080002), European Research Council (340755)), Puerto Rican Alzheimer Disease Initiative (PRADI) (RF1AG054074), Reasons for Geographic and Racial Differences in Stroke (REGARDS) (U01NS041588), Research in African American Alzheimer Disease Initiative (REAAADI) (U01AG052410), the Religious Orders Study (ROS) (P30 AG10161, P30 AG72975, R01 AG15819, R01 AG42210), the RUSH Memory and Aging Project (MAP) (R01 AG017917, R01 AG42210Stanford Extreme Phenotypes in AD (R01AG060747), University of Miami Brain Endowment Bank (MBB), University of Miami/Case Western/North Carolina A&T African American (UM/CASE/NCAT) (U01AG052410, R01AG028786), and Wisconsin Registry for Alzheimer's Prevention (WRAP) (R01AG027161 and R01AG054047).*

*The four LSACs are: the Human Genome Sequencing Center at the Baylor College of Medicine (U54 HG003273), the Broad Institute Genome Center (U54HG003067), The American Genome Center at the Uniformed Services University of the Health Sciences (U01AG057659), and the Washington University Genome Institute (U54HG003079).* *Genotyping and sequencing for the ADSP FUS is also conducted at John P. Hussman Institute for Human Genomics (HIHG) Center for Genome Technology (CGT).*

*Biological samples and associated phenotypic data used in primary data analyses were stored at Study Investigators institutions, and at the National Centralized Repository for Alzheimer's Disease and Related Dementias (NCRAD, U24AG021886) at Indiana University funded by NIA. Associated Phenotypic Data used in primary and secondary data analyses were provided by Study Investigators, the NIA funded Alzheimer's Disease Centers (ADCs), and the National Alzheimer's Coordinating Center (NACC, U24AG072122) and the National Institute on Aging Genetics of Alzheimer's Disease Data Storage Site (NIAGADS, U24AG041689) at the University of Pennsylvania, funded by NIA. Harmonized phenotypes were provided by the ADSP Phenotype Harmonization Consortium (ADSP-PHC), funded by NIA (U24 AG074855, U01 AG068057 and R01 AG059716) and Ultrascale Machine Learning to Empower Discovery in Alzheimer's Disease Biobanks (AI4AD, U01 AG068057). This research was supported in part by the Intramural Research Program of the National Institutes of health, National Library of Medicine. Contributors to the Genetic Analysis Data included Study Investigators on projects that were individually funded by NIA, and other NIH institutes, and by private U.S. organizations, or foreign governmental or nongovernmental organizations.*

### SUPPLEMENTAL REFERENCES

Adzhubei IA, Schmidt S, Peshkin L, Ramensky VE, Gerasimova A, Bork P, Kondrashov AS, Sunyaev SR. 2010. A method and server for predicting damaging missense mutations. *Nat Methods* **7**: 248–249.

Chen-Plotkin AS, Martinez-Lage M, Sleiman PMA, Hu W, Greene R, Wood EM, Bing S, Grossman M, Schellenberg GD, Hatanpaa KJ, et al. 2011. Genetic and clinical features of progranulin-associated frontotemporal lobar degeneration. *Arch Neurol* **68**: 488–497.

Cruchaga C, Haller G, Chakraverty S, Mayo K, Vallania FLM, Mitra RD, Faber K, Williamson J, Bird T, Diaz-Arrastia R, et al. 2012. Rare variants in APP, PSEN1 and PSEN2 increase risk for AD in late-onset Alzheimer's disease families. *PLoS One* **7**: e31039.

Kircher M, Witten DM, Jain P, O'Roak BJ, Cooper GM, Shendure J. 2014. A general framework for estimating the relative pathogenicity of human genetic variants. *Nat Genet* **46**: 310–315.

Mukherjee O, Wang J, Gitcho M, Chakraverty S, Taylor-Reinwald L, Shears S, Kauwe JSK, Norton J, Levitch D, Bigio EH, et al. 2008. Molecular characterization of novel progranulin (GRN) mutations in frontotemporal dementia. *Hum Mutat* **29**: 512–521.

Ng PC, Henikoff S. 2003. SIFT: Predicting amino acid changes that affect protein function. *Nucleic Acids Res* **31**: 3812–3814.

Shankaran SS, Capell A, Hruscha AT, Fellerer K, Neumann M, Schmid B, Haass C. 2008. Missense mutations in the progranulin gene linked to frontotemporal lobar degeneration with ubiquitin-immunoreactive inclusions reduce progranulin production and secretion. *J Biol Chem* **283**: 1744–1753.

Wallon D, Rousseau S, Rovelet-Lecrux A, Quillard-Muraine M, Guyant-Maréchal L, Martinaud O, Pariente J, Puel M, Rollin-Sillaire A, Pasquier F, et al. 2012. The French series of autosomal dominant early onset Alzheimer's disease cases: mutation spectrum and cerebrospinal fluid biomarkers. *J Alzheimers Dis* **30**: 847–856.
